# Evidence of shared genetic factors in the aetiology of gastrointestinal disorders and endometriosis and clinical implications for disease management

**DOI:** 10.1101/2022.10.20.22281201

**Authors:** Fei Yang, Yeda Wu, Richard Hockey, Jenny Doust, Gita D. Mishra, Grant W. Montgomery, the International Endometriosis Genetics Consortium, Sally Mortlock

## Abstract

In clinical practice, the co-existence of endometriosis and gastrointestinal symptoms is often observed; however, the factors driving this link remain largely unknown. Here, using large-scale multifaceted data including observational, genetic, and pharmaceutical datasets, we report a positive phenotypic and genetic association of endometriosis with peptic ulcer disease (PUD), gastro-oesophageal reflux disease (GORD), a combined GORD/PUD Medicated (GPM) phenotype and irritable bowel syndrome (IBS), but not with inflammatory bowel disease (IBD). Mendelian randomization analysis identified a causal effect of the GPM phenotype on endometriosis and a bidirectional causal association between endometriosis and IBS. Cross-trait meta-analysis and colocalization along with comprehensive functional annotation confirmed two shared genetic loci (*FN1, TACSTD2*) for endometriosis with IBS and twelve loci (*ETAA1, HOXC4, RERG, SEMA3F, SPAG16, HIST1H2BC, RAB5B, CCKBR* and *PDE4B*) with GORD and PUD. Shared genetic loci may contribute to risk of both endometriosis and digestive disorders through the involvement of DNA damage, estrogen regulated cell-proliferation and inflammation, and barrier dysfunction. Analyses of medication usage identified a higher use of drugs for IBS, GORD and PUD in women diagnosed with endometriosis as well as a higher use of hormone therapies in women diagnosed with IBS, GORD and PUD but not for IBD, which strongly supports the co-occurrence of these conditions and highlights the potential for drug repositioning and caution around drug contraindications in clinical practice. Taken together, the combined evidence robustly suggests a shared disease aetiology and provides important clinical implications for diagnostic and treatment decisions for endometriosis and digestive disorders.

**WHAT IS ALREADY KNOWN ON THIS TOPIC?:**

- Both endometriosis and gastrointestinal disorders affect a large proportion of people worldwide and the co-existence of endometriosis and gastrointestinal symptoms (eg, abnormal pain, bloating, constipation) is often observed in clinical practice.
- The association of these two diseases was supported but also limited to previous observational evidence which highlights a three-fold increase in the prevalence of irritable bowel syndrome (IBS) in women with endometriosis.
- Observational study is easily subject to measurement error, confounding and reverse causation. Therefore, it is important to assess the association using multidimensional datasets and more accurate approaches, such as the use of genetic data in a mendelian randomisation framework as well as the analysis from a perspective of medication usage.

**WHAT THIS STUDY ADDS:**

- Genetic risk factors for endometriosis and gastrointestinal disorders, two leading causes of discomfort and chronic pelvic pain, are correlated.
- Mendelian randomisation analyses supported a causal relationship between genetic predisposition to gastrointestinal disorders (gastro-oesophageal reflux disease (GORD) and peptic ulcer diseases (PUD)) and endometriosis risk, and evidence for a bidirectional causal relationship between endometriosis and IBS, which might explain in part the co-occurrence of these diseases.
- The identification of shared risk loci highlighted biological pathways that may contribute to the pathogenesis of both diseases, including estrogen regulation and inflammation, as well as potential therapeutic drug targets such as *CCKBR* and *PDE4B*.
- The higher use of drugs for IBS, GORD and PUD in women diagnosed with endometriosis as well as the higher use of hormone therapies in women diagnosed with IBS, GORD and PUD, support the co-occurrence of these conditions and shared disease aetiology but also highlights the potential for drug repositioning and caution around drug contraindications.

**Graphical abstract:** 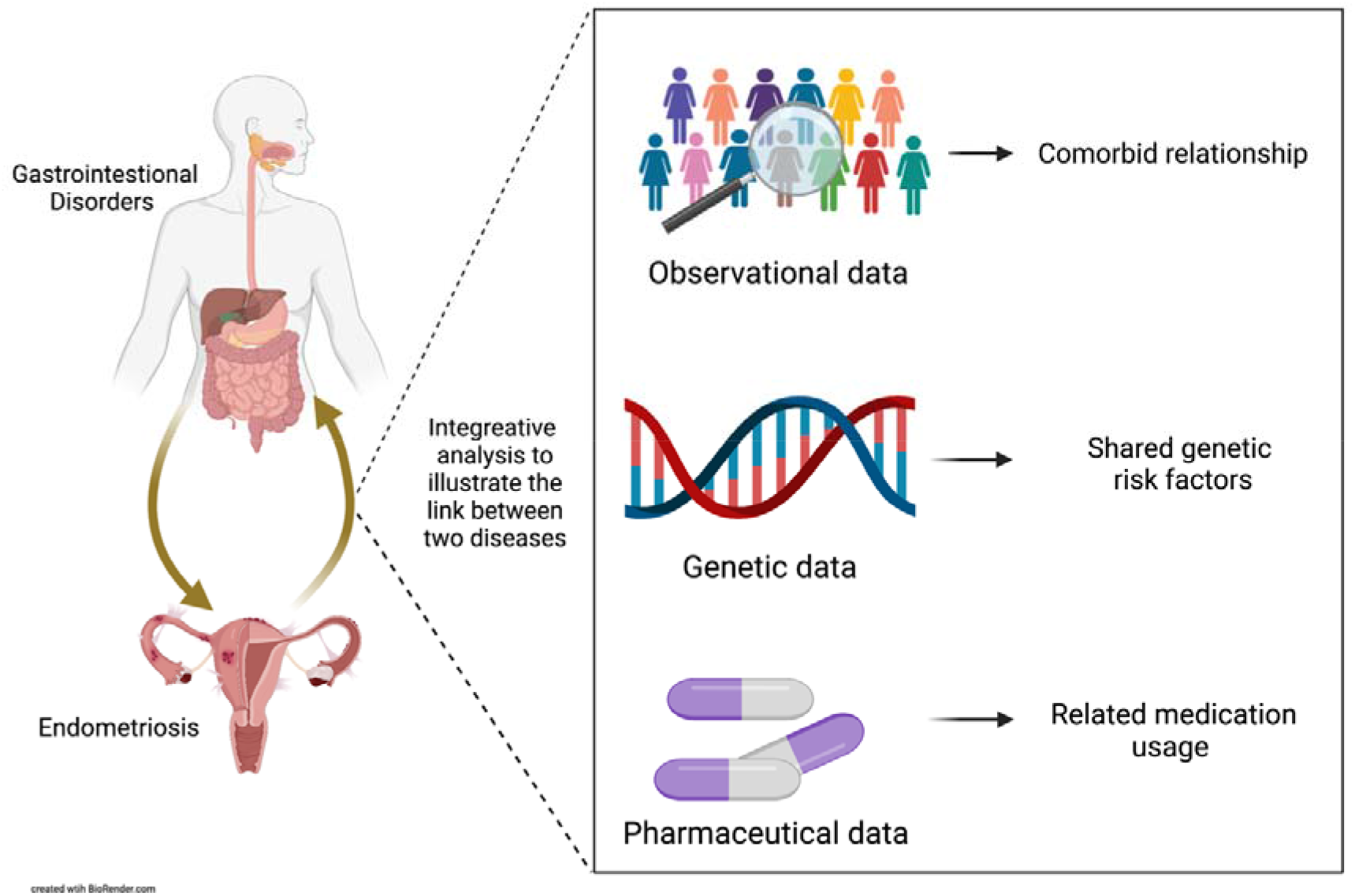

## Introduction

Endometriosis is a common gynaecological disease affecting around 11% of reproductive aged women, significantly impacting quality of life and work productivity^1 2^. The clinical manifestations of endometriosis are diverse. Many of the symptoms are non-specific, which preclude timely diagnosis and further prognosis^1 3^. It has often been observed that many women diagnosed with endometriosis also experience symptoms associated with gastrointestinal disorders (GI) including abdominal pain, bloating, constipation, heartburn, dyspepsia, vomiting, painful bowel movements, diarrhea, and nausea^4-6^. Studies have shown that whilst these symptoms do not necessarily involve bowel lesions associated with endometriosis, symptoms such as cyclic-related bloating, constipation and diarrhea can get worse during menstruation^5-8^. Inevitably, this presents challenges for clinicians to accurately diagnosis both diseases among women. Understanding shared disease aetiology critically impacts both disease diagnosis and management.

Previous observational studies have provided some evidence for the associations between endometriosis and digestive disorders. Meta-analyses have reported a three-fold increase in the prevalence of IBS in women with endometriosis compared to women without endometriosis^9 10^. This was supported by a recent retrospective study using a large nationwide biobank-based cohort, the Estonian Biobank (EstBB), that reported a notable proportion of women diagnosed with endometriosis or IBS also suffered from IBS (13.6%) or endometriosis (9.0%), respectively^11^. A nationwide Danish cohort study found a significantly increased risk of inflammatory bowel disease (IBD) in endometriosis patients with a standardised incidence ratio (SIR) of 1.5 (95% confidence interval 1.4 to 1.7), and the relationship became stronger when restricted to surgically confirmed endometriosis^12^. Currently, very few observational studies have investigated the association between endometriosis and other gastrointestinal disorders. However, endometriosis symptoms overlap with other common gastrointestinal disorders, including peptic ulcer disease (PUD) and gastro-oesophageal reflux disease (GORD)^13^.

Despite studies showing endometriosis patients are more likely than people without endometriosis to present gastrointestinal symptoms or have a diagnosis of gastrointestinal disorders, it is uncertain whether this is due to 1) a direct effect of endometriosis itself; 2) shared aetiological factors between endometriosis and gastrointestinal diseases; 3) side effects of medical treatments; or 4) the inevitable association bias in observational studies, such as the measurement error, reverse causation, residual or unmeasured confounding^14^. For example, therapeutic use of gonadotropin-releasing hormone (GnRH) analogues and nonsteroidal anti-inflammatory drugs (NSAIDs), to manage symptoms of endometriosis, has been widely reported to aggravate the severity of gastrointestinal symptoms and contribute to gastrointestinal disorders including PUD^6 15-18^. Therefore, using a more accurate approach and multidimensional dataset to validate the relationship between endometriosis and gastrointestinal disorders has important implications, not only for understanding shared disease mechanisms but also for informing therapeutic strategies in clinical practice.

Endometriosis and gastrointestinal diseases mentioned above are common multifactorial diseases with environmental and genetic risk factors both playing roles in the development of these diseases^19 20^. Twin and family studies have shown a heritable component of endometriosis, IBS, PUD, IBD and GORD^21-26^. Genome-wide association study (GWAS), which have been widely used to identify disease associated genetic risk variants^27^, also enable the use of mendelian randomisation (MR) approaches to assess the causal effect of cumulative genetic predisposition to one disease on other disease risk. Given the advantage that genetic alleles are randomly determined at conception and free from potential environmental confounders (eg. medication usage) and measurement error due to self-report, MR analysis can minimize the biases that frequently weaken results obtained from observational approachs^14 28^. Therefore, application of GWAS data and an MR framework is of great value in understanding shared aetiology.

One study recently implicated causal links between endometriosis and depression with gastric mucosa abnormalities using genetic data^29^. However, the relationship between endometriosis and other common GI disorders, like IBS and IBD and PUD, were not investigated. GWAS studies have identified risk variants for endometriosis and gastrointestinal disorders independently^30-32^, including one female-specific IBS risk locus at 9q31.2, which was also previously reported as more strongly associated with early age at menarche^32^, a risk factor for endometriosis. This study presents a comprehensive evaluation of the relationship between endometriosis and gastrointestinal disorders through analysis of large-scale genetic datasets, and epidemiological and pharmaceutical data in the UK Biobank (UKB) and Australian Longitudinal Study on Women’s Health (ALSWH)^33 34^.

## Method

### Data resources

The large-scale GWAS summary statistics for endometriosis and five gastrointestinal disorder phenotypes, utilized in this study, have been well described in previous studies^30 31^. Summary data for endometriosis were restricted to eight European ancestry cohorts^31 35^ for the purpose of this analysis. A total number of 14,926 cases and 189,715 controls genotyped across 7,899,415 SNPs were included in the endometriosis meta-analysis. GWAS summary statistics for four gastrointestinal disorders, gastro-oesophageal reflux disease (GORD) (n_case_ = 39,851, n_control_ = 416,563), peptic ulcer disease (PUD) (n_case_ = 12,226, n_control_ = 444,188), irritable bowel syndrome (IBS) (n_case_ = 14994, n_control_ = 441420) and inflammatory bowel disease (IBD) (n_case_ = 6,115, n_control_ = 450,299), were previously generated using genetic data from individuals in the UK Biobank (UKB), using the health-related outcomes data from combing self-reported, primary care, death register and hospital reported diagnoses. As medications used for PUD also have a therapeutic effect on GORD, a fifth phenotype for gastrointestinal disorders is the combined GORD and PUD and individuals taking medications for GORD/PUD making a total of 75,192 cases and 381,222 controls in the GORD/PUD Medicated (GPM) phenotype^30^. Sex stratified GWAS summary statistics for IBS were also generated in this study for the purpose of exploring potential sex bias in the relationship between endometriosis and IBS.

### Comorbidity analysis

As a cross-sectional analysis, the comorbid relationship between endometriosis and each gastrointestinal disorder (IBS, IBD, GORD and PUD) described above were investigated among unrelated European female individuals in the UKB, with ancestry definition described previously^36^. Phenotypes were defined using self-reported, hospital admission, death register or primary care record data. Endometriosis cases were defined using date and source of endometriosis reported (UKB data fields:132122 & 132123), ICD10 diagnosis (UKB data field:41270), ICD9 diagnosis (UKB data field: 41271) and self-report (UKB data field: 20002), totalling 5,392 cases (excluding endometriosis of the uterus/adenomyosis). Gastrointestinal disorders definitions were similar as previously described by Wu et al.^30^ however, these were also restricted to females. A total of 16,330 IBS cases were included (UKB data field: 131639) alongside 22,383 GORD cases (UKB data field: 131585). IBD and PUD were defined using a combination of disease codes, IBD cases were a combination of Crohn’s diseases (UKB data field: 131627) and ulcerative colitis (UKB data field: 131629) diagnoses totalling 2,708 cases and PUD cases were a combination of gastric ulcer cases (UKB data field : 131591), duodenal ulcer cases (UKB data field: 131593), other site peptic ulcer cases (UKB data field: 131595) and gastro-jejunal ulcer cases (UKB data field : 131597) totalling 5,208. We firstly measured whether individuals diagnosed with endometriosis were more likely to have a diagnosis of IBS, IBD, GORD and/or PUD using Fisher’s exact test. Next, we conducted a competitive comorbidity analysis to test which digestive disorder is more prone to be comorbid with endometriosis among these four disorders. Briefly, the proportion of endometriosis cases in each of the four digestive diseases were calculated and then compared in pairs using a two-proportion Z-test. To meet the prerequisite of this analysis that samples in each pair are independent, we removed overlapping samples when calculating the proportion.

### Genetic correlation

Genetic correlation attributable to the genome-wide common SNPs between endometriosis and each of the other five gastrointestinal disorder phenotypes (IBS, IBD, GORD, PUD, GPM) was estimated using bivariate linkage disequilibrium score regression (LDSC)^37^ and their respective GWAS summary statistics. GWAS summary data was formatted using the function ‘munge_sumstats.py’ outlined in the LDSC manual and the genetic correlation for each pair was estimated. The European 1000 genome reference data was adopted in the calculation of linkage disequilibrium (LD) scores. A sex-stratified analysis was conducted to further investigate whether the genetic correlation between endometriosis and IBS is sex dependent. Given that there was no sample overlap in GWAS studies of endometriosis and gastrointestinal disorders and all participants are of European ancestry, we also reduced standard error of genetic correlations by constraining the intercept, which was used to protect bias from population stratification and sample overlap in different GWAS studies.

### Assessing potential causal relationships

Mendelian randomisation (MR) uses genetic variants that are robustly associated with exposure of interest to test whether those genetic variants also increase the risk of another trait ^14^. MR has emerged as a valuable tool to assess the causal effect of one trait on another. The genetic variants selected are robust and are not associated with other confounders and will only influence the outcome trait through the trait of interest if there is a causal association, thus less susceptible to confounding, measurement error, and reverse causation when compared with conventional observation studies^14 38 39^. In this study, the causal relationships between endometriosis and gastrointestinal disorder phenotypes (IBS, GORD, PUD, GPM) was investigated using one wildly-accepted MR method called Generalised Summary-data-based Mendelian randomization (GSMR)^40^. The combined phenotype of GPM was used in place of individual GWAS for PUD and GORD to increase study power. GSMR uses all significantly associated SNPs as SNP instruments to test for causality. To reduce the influence of horizontal pleiotropy (a single locus directly affecting multiple phenotypes), one potential confounding factor for Mendelian randomization analysis, we also applied the HEIDI-outlier analysis to detect SNPs having obvious pleotropic effect on both risk factor and diseases. To remove potential confounding from the correlation between the GI traits, we applied mtCOJO^40^ on both the exposure and outcome trait. We further used the adjusted GWAS summary statistics to repeat the GSMR analysis again. A *P*-value of < 0.05 was considered significant. In some cases there was an insufficient number of SNPs to use as instruments and so the GWAS threshold was relaxed to allow at least ten SNPs for each phenotype, following the author’s recommendation to include at least 10 SNP instruments during GSMR analysis to achieve robust results.

### Cross-trait meta-analysis of endometriosis and gastrointestinal diseases

We next adopted two complementary cross-trait meta-analysis methods, MetABF^41^ and Eskin random-effects model (RE2C)^42^, to identify whether there are shared risk loci between endometriosis and the digestive disorders (IBS, GPM), as well as potential novel risk loci for each disease. MetABF performs the multi-trait meta-analysis based on the Bayesian framework. Effect alleles were harmonised across all three GWAS. Both fixed and independent effect models were used when performing this meta-analysis. The prior parameter accounting for effects of heterogeneity in two diseases was set as 0.1, which is typically used in complex diseases. As a result, SNPs with a logABF > 4 and at least a normally significant *P*-value < 0.05 in each individual disease GWAS analysis were defined as significant in the MetABF analysis.

To validate the MetABF results, we used a complementary cross-trait meta-analysis approach, RE2C, which dramatically increases power when statistics among different studies are correlated compared with other methods. RE2C also accounts for the heterogeneous effects within studies using a novel statistic model. Similar to MetABF, effect alleles were harmonised across the GWAS prior to being used as input for the RE2C analysis. As a result, a SNP meeting the *P*-value threshold of < 5e-8 in either fixed (Lin-Sullivan method) or random (RE2C) effects model and having at least a normally significant *P*-value < 0.05 in the individual disease GWAS, were deemed as significant in the meta-analysis. SNPs meeting both thresholds of MetABF and RE2C were selected for the further fine mapping analysis in Functional Mapping and Annotation (FUMA)^43^ to identify independent risk loci using a threshold of *r*^2^ < 0.6 and then the lead SNPs at a threshold of *r*^2^ < 0.1. The maximum distance between LD blocks to merge into a locus were set as 250kb.

### Colocalization analysis

To identify specific genomic regions that have the same causal variant for each disease we conducted a pairwise GWAS (GWAS-PW)^44^ analysis. Again, this analysis was restricted to comparisons with IBS and GPM. The input to GWAS-PW is a set of estimated effect sizes and standard error for each SNP on each of the paired diseases. The whole genome was split into 1,703 LD independent blocks, and the probability is estimated for four models extended from Giambartolomei et al^45^, that a given region (a) contains a genetic variant that impacts first disease (PPA1); (b) contains a genetic variant that impacts the second disease (PPA2); (c) contains a genetic variant that affects both diseases; (PPA3) or (d) contains two distinct variants that influences each disease separately (PPA4). Paired summary statistics for endometriosis with IBS or GPM were analysed. Any regions that were identified with a PPA3/PPA4 >0.5 were considered to show evidence of a shared causal variant and two distinct causal variants respectively.

### Functional annotation and gene mapping

In order to identify potential target genes associated with both endometriosis and IBS or GPM, we used FUMA to perform Multi-marker Analysis of GenoMic Annotation (MAGMA) gene-set analysis and two additional annotation approaches, positional mapping with combined annotation-dependent depletion (CADD) score and cis-expression quantitative trait loci (eQTL) mapping for independent SNPs and SNPs in LD identified in the above cross-trait meta-analysis. Gene sets were adopted from Genotype-Tissue Expression (GTEx) project which collects 54 non-diseased tissue sites across nearly 1000 individuals^46^. Cis-eQTL information of 12 digestive and reproductive tissues (Supplementary Table 1) in the GTEx Project^46^, endometrium eQTLs^47^ and a large blood eQTL dataset, eQTLGen^48^ were used for the eQTL mapping analysis. Variants with a CADD score of more than 12.7 were defined as potentially pathogenic.

SMR analysis is a powerful approach to identify likely causal relationship between the trait-associated SNPs and gene expression. SMR analysis was performed on endometriosis, IBS and GPM respectively using eQTL data from 12 digestive and reproductive tissues in the GTEx project^46^ and endometrium^47^. Associations were defined as significant if *P*_SMR_ < 0.05 and *P*_HEIDI_ > 0.05. We determined if any SMR significant genes were shared between the diseases. Shared causal associations indicated that SNPs may be associated with both diseases through the regulation of expression of the same gene.

Additional epigenomic functional annotation was performed with EpiMap^49^ using epigenome maps from relevant tissues (uterus, ovary, gastroesophageal sphincter, Peyer’s patch, oesophagus, stomach, colon, intestine, rectum) to define chromatin states, enhancers, upstream regulators and downstream target genes.

### Pathway-based enrichment analysis

To identify which biological pathways are associated with both endometriosis and IBS or GPM, we firstly performed a gene-set enrichment analysis using MAGMA implemented in FUMA. Significant SNPs from the cross-trait meta-analysis were used as input for MAGMA and a window of 0kb outside of a gene was adopted in the gene-set analysis. In addition, genes annotated to significant loci from the cross-trait meta-analysis were included in a GENE2FUNCTION analysis in FUMA to identify whether these genes were enriched in any curated gene sets.

### Phenome-wide association

In order to investigate whether the correlation between endometriosis and each of IBS and GPM can be explained by the genetic susceptibility to any other traits or diseases, we searched traits that were associated with genome-wide significant independent SNPs and SNPs in LD (r^2^ > 0.8) from the cross-trait meta-analysis. We used information from GWAS catalogue and PhenoScanner ^50^.

### Drug target analysis

Using the online Open-targets drug database (www.targetvalidation.org), we investigated if any known drug targets are common across endometriosis and gastrointestinal disorders (GORD, PUD, IBS) and if any genes functionally annotated to shared risk loci are potential drug targets for either endometriosis and/or digestive disorders.

### Medication usage

To investigate the implications of medication use on the relationship between endometriosis and gastrointestinal disorders we analysed Pharmaceutical Benefits Scheme (PBS) data by endometriosis status from the 1973-78 and 1989-95 cohorts in the Australian Longitudinal Study on Women’s Health (ALSWH)^33 34^. The frequency of medications used in these women was calculated. In addition, we characterized the medication use (UKB data field: 20003) in unrelated women in the UK Biobank, including 5,392 women diagnosed with endometriosis, 15,881 women with IBS, 22,383 women with GORD, 5,208 women with PUD and 2,708 women with IBD. We randomly selected age-matched controls to avoid the potential bias caused by differences in age distribution between cases and controls. Differences in the proportion of women, with and without an above diagnosis, using reported medications was tested using fisher test. After correcting for multiple testing using Bonferroni analysis, a *P*-value of < 0.05 was considered significant.

## Results

### Significant comorbid relationship between endometriosis and GI disorders

Using data from 188,461 unrelated females in UKB, we found a bidirectional epidemiological relationship between endometriosis and each of the four GI disorders (IBS, IBD, GORD, PUD). GORD and IBS remained significant after accounting for multiple tests. Of those associations (Table 1), women with endometriosis were two times more likely to have an IBS diagnosis (OR = 2.01, 95% CI 1.86 to 2.16; P-value = 3.90e-68), and 1.4 times more likely to have a GORD diagnosis (OR = 1.40, 95% CI 1.30 to 1.50; p = 3.54e-18), than those free of endometriosis. The competitive comorbidity analyses confirmed that women with endometriosis are more prone to be comorbid with IBS, followed by GORD, when compared with PUD (OR = 1.22, 95% CI 1.05 to 1.42; p = 0.01) and IBD (OR = 1.25, 95% CI 1.01 to 1.53; p = 0.04) (Figure 1).

**Table 1.** Comorbid relationship between endometriosis and gastrointestinal disorders (GI) in unrelated European women in the UK Biobank. Association between diseases was tested using the fisher’s exact test.

|  | GORD<br>(n=22,383) | IBS<br>(n=16,330) | PUD<br>(n=5,208) | IBD<br>(n=2,708) |
| --- | --- | --- | --- | --- |
| **Endo vs GI** | 1.40 (1.30-1.50)<br>*P*=3.54e-18 | 2.01 (1.86-2.15)<br>*P*=3.90e-68 | 1.22 (1.05-1.42)<br>*P*=0.01 | 1.25 (1.01-1.53)<br>*P*=0.04 |
| **GI vs Endo** | 1.45 (1.35-1.57)<br>*P*=5.94e-22 | 2.13 (1.98-2.30)<br>*P*=7.14e-79 | 1.22 (1.05-1.42)<br>*P*=0.01 | 1.24 (1.0-1.53)<br>*P*=0.04 |

Note: GORD (Gastro-oesophageal Reflux Disease), IBS (Irritable Bowel Disease), PUD (Peptic Ulcer Disease) and IBD (Inflammatory bowel disease), Endo (Endometriosis). The number in the bracket below each disease represent the number of women diagnosed with this disease. There are a total number of 5,392 women diagnosed with endometriosis. Fisher's exact results are represented by OR, 95% CI values within brackets and P-value.

**Figure 1.**
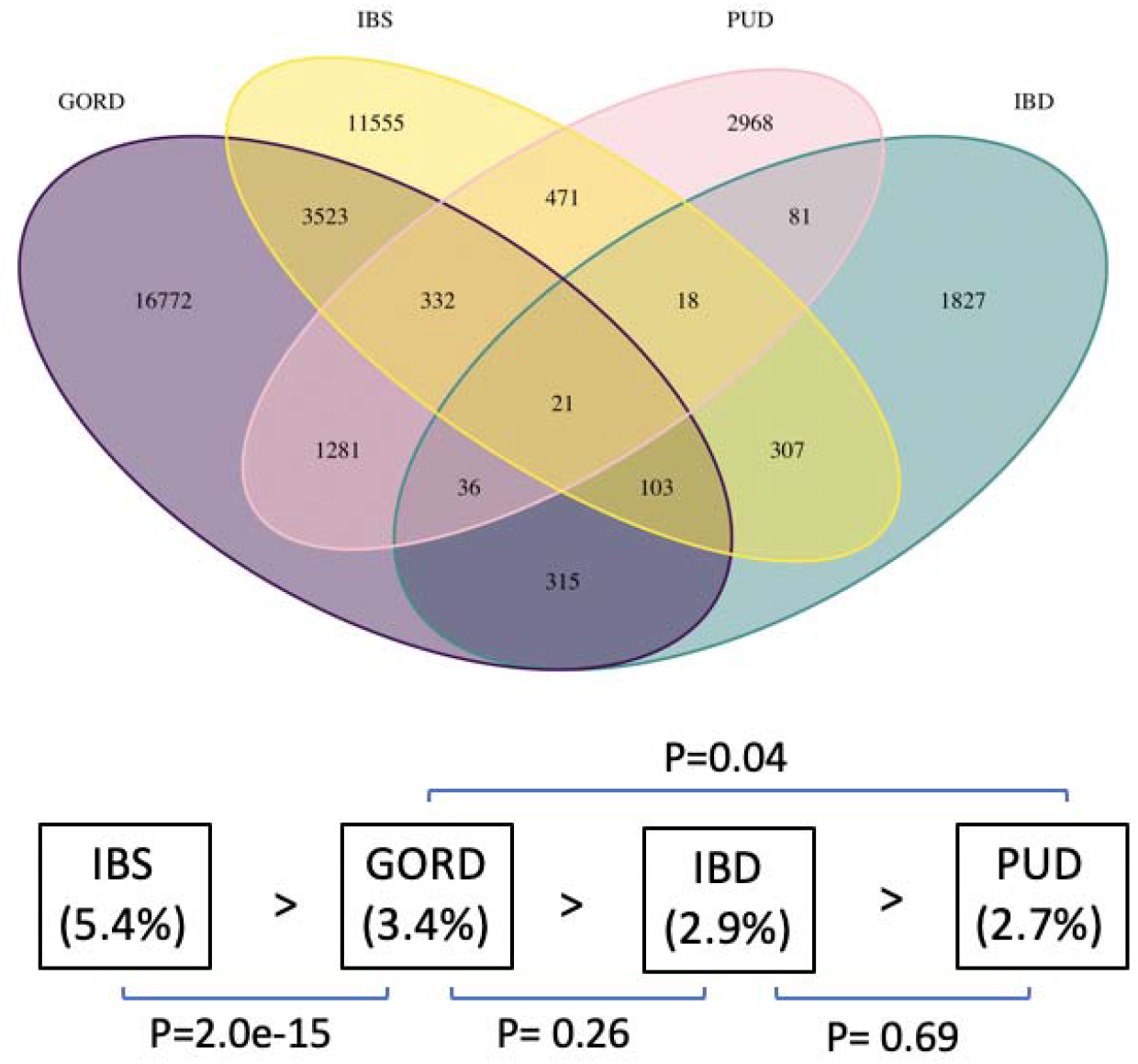
Competitive comorbidity analysis for endometriosis and four gastrointestinal disorders in unrelated female European individuals. The Venn diagram shows the number of individuals with diagnosis of gastrointestinal disorders. At the bottom of Venn diagram is the proportion of endometriosis cases (n=5,392) in each of the digestive diseases after removing the overlapped individuals for these four diseases, ranking from highest proportion to the least. Paired comparison was conducted using a two-proportion Z test, with the corresponding *P*-value under each comparison.

### Genetic correlation between endometriosis and GI disorders

Compared with comorbidity relationship identified above, the LDSC analysis only provided evidence of a significant positive genetic correlation (rg) between endometriosis and IBS (rg = 0.22, p = 0.005), GORD (rg = 0.16, p = 0.004), PUD (rg = 0.23, p = 0.003) and GPM (rg = 0.22, p = 2.17e-06) (Figure 2A). There was no evidence of significant correlation between endometriosis and IBD. Constraining the intercept due to no sample overlap, resulted in a smaller standard error whereas there was little change in the genetic correlation (Figure 2A).

**Figure 2.**
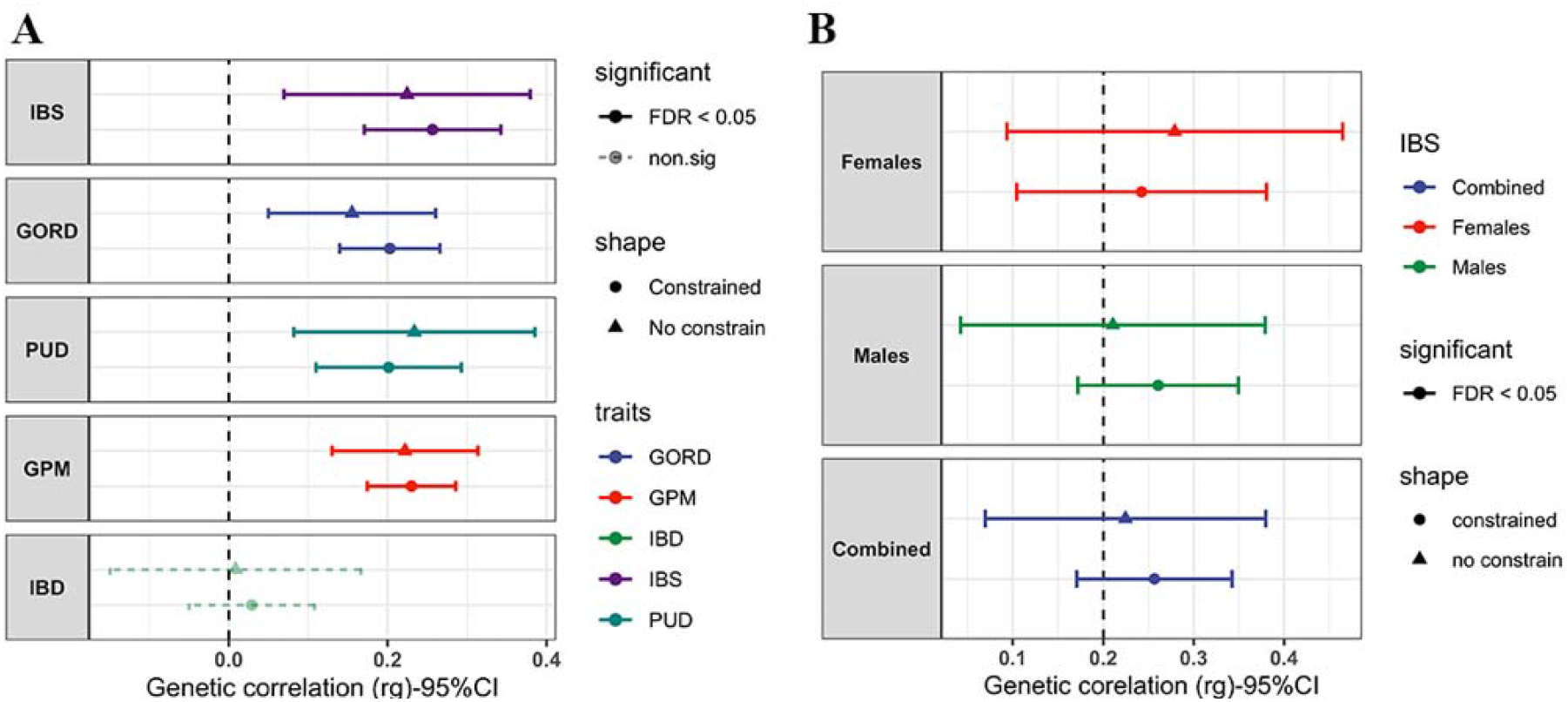
Results of genetic correlation between endometriosis and a) gastrointestinal disorders (irritable bowel syndrome (IBS), peptic ulcer disease (PUD), gastro-oesophageal reflux disease (GORD), GORD/PUD Medicated (GPM) inflammatory bowel disease (IBD)) and b) separate male and female IBS GWAS datasets. The x-axis indicates the value of genetic correlation, and the error bar indicates its 95% confidence interval. All red lines represent the results after constraining the heritability intercept to one considering no sample overlap for each comparison.

Although endometriosis is rarely observed in males, males do carry endometriosis risk alleles, as such we also conducted the genetic correlation analysis separately for females and males. The results remained significant between endometriosis and both the separate female (rg = 0.28, p = 0.003) and male (rg = 0.21, p = 0.014) IBS GWAS cohorts (Figure 2B).

### Complex causal relationship between endometriosis and GI disorders

Following the identification of shared genetic correlation, we applied an MR method^40^ to estimate the causal relationship between GPM, IBS, and endometriosis (Figure 3). We identified evidence of a significant association between GPM and endometriosis whereby genetic variants contributing to the risk of GPM (genetic predisposition to GPM) also increased risk of endometriosis (odds ratio = 1.56 (95% CI 1.35 to 1.76), p = 2.47e-5) (Table 2). The reverse MR analysis (the genetic effect of endometriosis on GPM) was not statistically significant (odds ratio = 0.98 (95% CI 0.94 to 1.02), p = 0.32). Due to the limited number of SNP instruments available for IBS when using a genome-wide significant level (*P* < 5e-8), we were unable to estimate the effect of IBS on endometriosis risk. However, using endometriosis as the exposure we identified that genetic variants contributing to the risk of endometriosis had a small effect on risk to IBS (odds ratio=1.07 (95% CI 1.01 to 1.13), p = 0.042). This effect was no longer significant following stringent Bonferroni correction for multiple testing.

**Figure 3.**
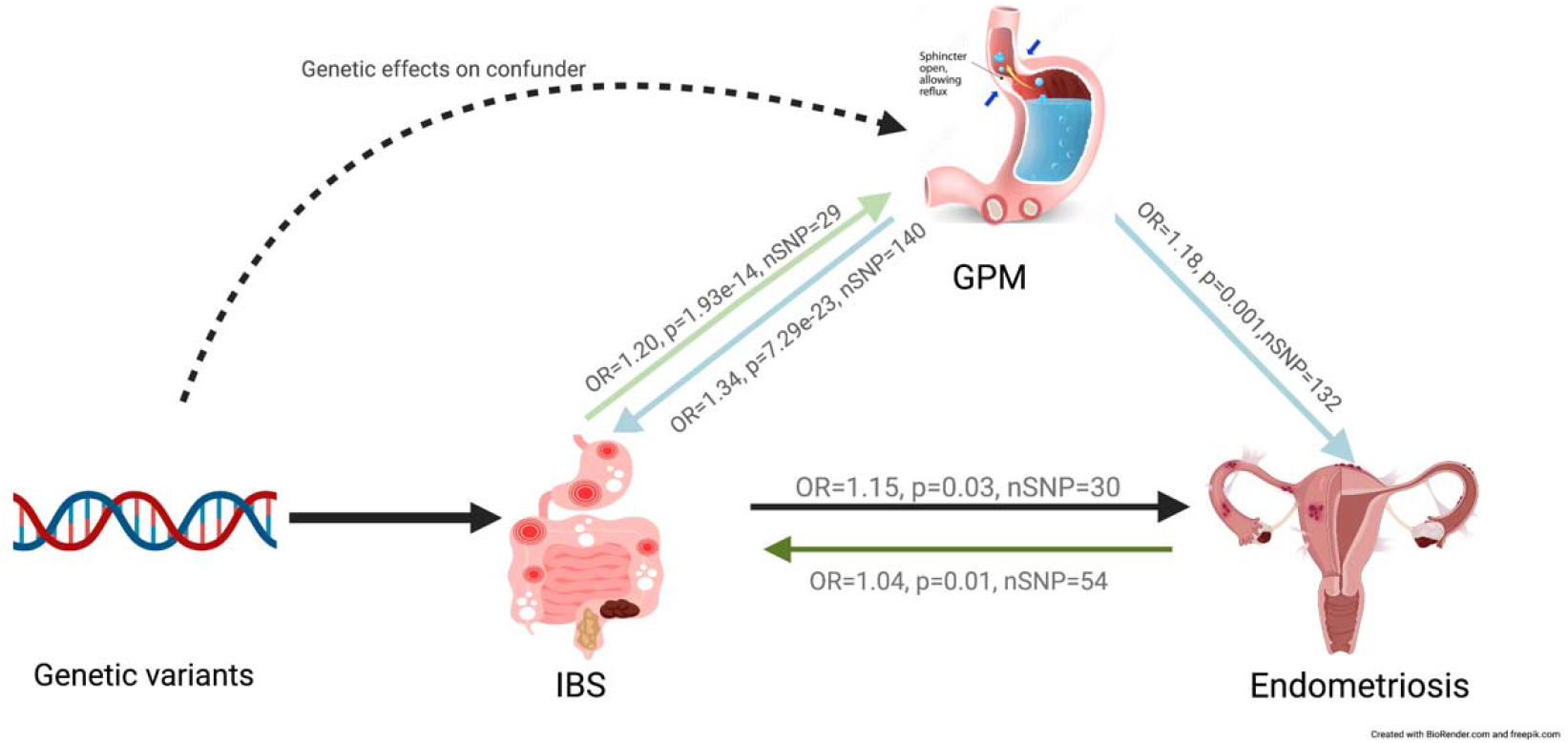
Simplified causal relationship identified by GSMR (Generalised Summary-data-based Mendelian Randomisation). Different arrow colour represents the specific direction of causal relationship.

**Table 2.** Summary of GSMR results for endometriosis (Endo), irritable bowel syndrome (IBS) and GORD/PUD Medicated (GPM) using different SNP instrument GWAS p-value thresholds.

| Exposure | Outcome | Odds Ratio<br>(95% CI) | SE | P-value | No. SNP<br>Instruments |
| --- | --- | --- | --- | --- | --- |
| **SNP instrument GWAS p-value<5e-8** |  |  |  |  |  |
| GPM | Endo | 1.56 (1.35, 1.76) | 0.104 | 2.47e-05 | 20 |
| GPM<sub>IBS</sub> | Endo<sub>IBS</sub> | 1.22 (1.00, 1.44) | 0.112 | 0.073 | 17 |
| Endo | IBS | 1.07 (1.01, 1.13) | 0.031 | 0.042 | 10 |
| Endo<sub>GPM</sub> | IBS<sub>GPM</sub> | 1.09 (1.03, 1.14) | 0.028 | 0.003 | 12 |
| **SNP instrument GWAS p-value<5e-6** |  |  |  |  |  |
| GPM | Endo | 1.18 (1.08, 1.27) | 0.049 | 0.001 | 132 |
| GPM<sub>IBS</sub> | Endo<sub>IBS</sub> | 1.16 (1.05, 1.27) | 0.055 | 0.008 | 103 |
| Endo | IBS | 1.04 (1.01, 1.07) | 0.016 | 0.012 | 54 |
| Endo<sub>GPM</sub> | IBS<sub>GPM</sub> | 1.03 (1.00, 1.06) | 0.016 | 0.050 | 48 |
| IBS | Endo | 1.15 (1.03, 1.28) | 0.066 | 0.028 | 30 |
| IBS<sub>GPM</sub> | Endo<sub>GPM</sub> | 1.23 (1.09, 1.38) | 0.074 | 0.004 | 25 |
| <sub>IBS</sub> = primary trait conditioned on IBS using mtCOJO |  |  |  |  |  |
| <sub>GPM</sub> = primary trait conditioned on GPM using mtCOJO |  |  |  |  |  |
| 95% CI = 95% confidence interval |  |  |  |  |  |

To increase the number of SNP instruments in the analysis we relaxed the SNP inclusion threshold to *P* < 5e-6 and repeated the GSMR analysis. The relationship between GPM and endometriosis, and endometriosis and IBS, was consistent, although attenuated, compared with that estimated using the genome-wide significant level (*P* < 5e-8) (Table 2). Interestingly, we also found evidence of bi-directional relationship between IBS and endometriosis. Genetic variants that increased risk of IBS had a significant risk effect on endometriosis (odds ratio=1.15 (95% CI 1.03 to 1.28), p = 0.028) however, this effect was no longer significant following stringent Bonferroni correction for multiple testing.

A genetic correlation and strong bi-directional association between IBS and GPM has been previously reported^30^, with genetic predisposition to IBS having a causal effect on GPM (odds ratio = 1.20 (95% CI 1.15 to 1.24), p = 1.93e-14) and genetic predisposition to GPM having a causal effect on IBS (odds ratio = 1.34 (95% CI 1.28 to 1.40), p = 7.29e-23). The complex causal relationship among the three diseases is illustrated in Figure 3. The relationship between GI disorders may act as a confounder impacting the results of each pair of MR analyses. To avoid the potential effect of GPM on the relationship between endometriosis and IBS, we adjusted the GWAS data of both endometriosis and IBS for the effects of GPM using mtCOJO to identify disease specific variant associations independent of GPM. Following this conditional analysis, evidence for the bi-directional causal relationship between IBS and endometriosis remained (Table 2, Figure 4). The causal relationship between GPM and endometriosis, following adjustment for the genetic effects of IBS, was only significant when the less stringent SNP inclusion p-value threshold was used but did not pass correction for multiple testing (odds ratio = 1.16 (95% CI 1.05 to 1.27), p = 0.008) (Table 2, Figure 4).

**Figure 4.**
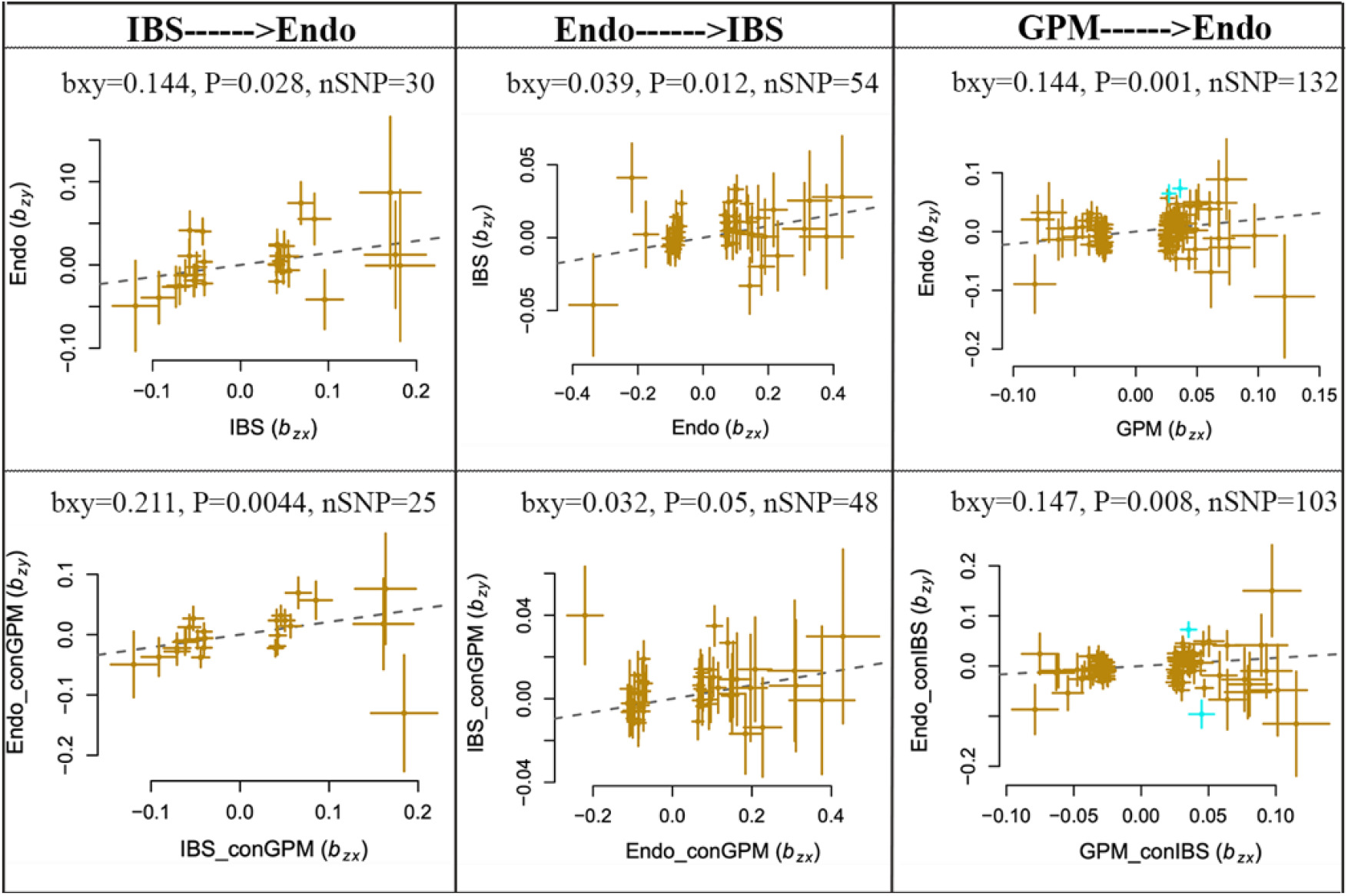
GSMR results for gastrointestinal disorders and endometriosis. The first row represents results using original GWAS data while the second row used the GWAS data conditioned on GPM or IBS. GWAS threshold for SNP instrument was set as *P* < 5e-6. bxy is the effect of exposure on outcome free of confounding from non-genetic factors and can be approximately interpreted as log(odds ratio).

MR was run separately for GORD and PUD to test if the relationship between GPM and endometriosis was driven by one particular phenotype. There was evidence of a significant relationship between GORD and endometriosis which remained when conditioning both traits on PUD (OR=1.15 (95% CI 1.10 to 1.21), p=1.69e-6) however, there was no evidence of a causal relationship between PUD and endometriosis (Supplementary Table 2). More powerful GWAS studies may be required to validate these causal relationships.

### Genomic loci associated with both endometriosis and GI disorders

To identify if there are any risk loci associated with both endometriosis and IBS or GPM, we carried out a cross-trait meta-analysis using two different methods, MetABF and RE2C. SNPs were considered associated with both diseases at a genome-wide significant level if they had a logABF > 4 in MetABF and a *P*-value < 5e-8 in RE2C models and a *P*-value < 0.05 in each individual GWAS analysis. As a result, a total number of 477 SNPs met criteria for endometriosis and GPM while only 32 SNPs were significant for endometriosis and IBS. Using FUMA, 12 genomic risk loci (21 independent signals) were identified as significantly associated with both endometriosis and GPM and three with endometriosis and IBS (Table 3). Among those loci identified by the cross-trait meta-analysis, the SNP on chr2:67845739 (rs2861694) within *ETAA1* was previously reported as associated with both endometriosis and GPM, another five SNPs were significantly associated with either endometriosis or GPM. The remaining nine risk loci were identified for the first time at a genome-wide level of significance for endometriosis and GPM and IBS (Table 3).

**Table 3.**
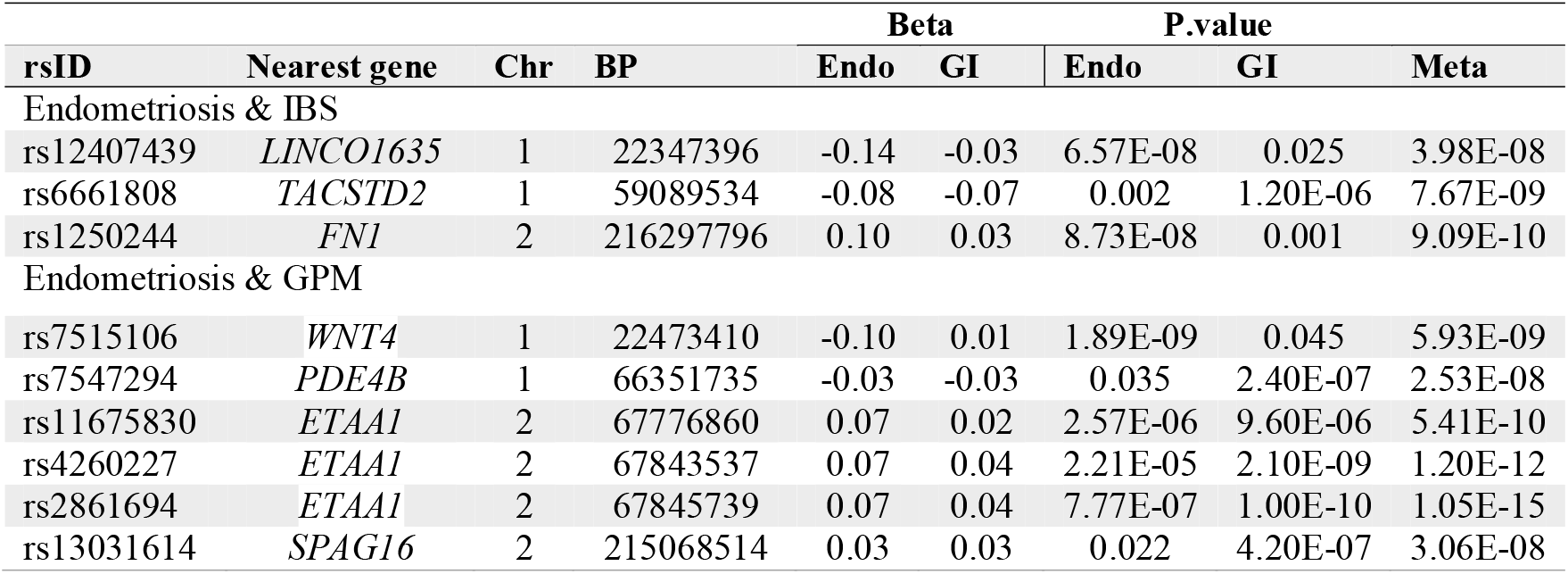
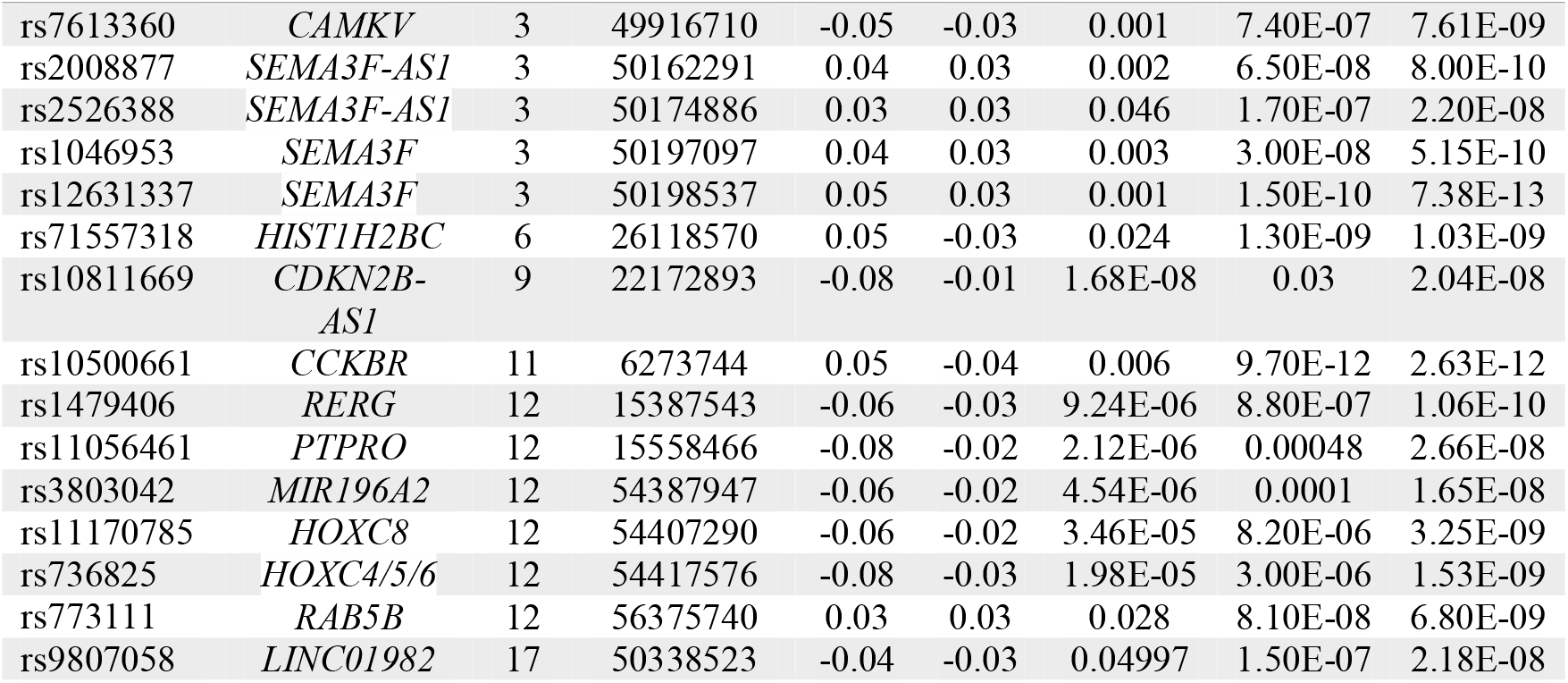
Significant SNP loci identified by endometriosis (Endo), irritable bowel syndrome (IBS) and GORD/PUD Medicated (GPM) cross-trait meta-analysis.

### Evidence of shared causal variants between endometriosis and GI disorders

GWAS-PW was used to perform a colocalization analysis to assess if any of the genomic regions jointly affected endometriosis and IBS. Results (Table 4) showed that there were three regions with PPA3 > 0.5 and three regions with PPA4 > 0.5, suggesting that 3 regions contain the same causal variant for both endometriosis and IBS whereas another three regions contain distinct and independent causal variants for endometriosis and IBS respectively. Analysing endometriosis and GPM, we identified six regions with PPA3 > 0.5 and 60 regions with PPA4 > 0.5. None of the regions with PPA3 > 0.5 in endometriosis and GPM overlapped with the three regions identified between endometriosis and IBS. Regions with evidence of a single casual variant (PPA3 > 0.5) for both endometriosis and either IBS or GPM are shown in Table 4. Among these identified regions, the two loci (*TACSTD2* and *FN1*) with the highest probability of a shared causal variant for endometriosis and IBS and the three loci *(ETAA1, HOXC4* and *RERG*) with the highest probability of a shared causal variant for endometriosis and GPM were also identified by the cross-trait meta-analysis described above. Specifically, the region with the strongest pleiotropic effect on endometriosis and GPM is located near *ETAA1/LINCO1812* on chromosome 2 (Figure 5), SNPs in this region are significantly associated with these two diseases and the index SNPs in the individual GWAS studies are in strong LD (r^2^ = 0.88). Other regions identified by both cross-trait meta-analysis and GWAS-PW are shown in Supplementary Figure 1A-G.

**Table 4.** Genomic regions that contain a same causal variant jointly influencing endometriosis with irritable bowel syndrome (IBS) and GORD/PUD Medicated (GPM) respectively.

|  |  |  |  |  | Risk loci (*P* < 5e-8) |  |  |
| --- | --- | --- | --- | --- | --- | --- | --- |
| Chr | Start | End | PPA-3 | Nearest gene | Endo | GI | Meta |
| Endometriosis & IBS |  |  |  |  |  |  |  |
| 2 | 215573795 | 217715180 | 0.818 | FN1 | Yes | No | Yes |
| 1 | 58865399 | 59889341 | 0.766 | OMA1/ TACSTD2/MYSM1 | No | No | Yes |
| 6 | 30798252 | 31565648 | 0.532 | HLA-C | No | No | No |
| Endometriosis & GPM |  |  |  |  |  |  |  |
| 2 | 67227143 | 68009259 | 0.983 | ETAA1/ LINC01812 | Yes | Yes | Yes |
| 12 | 53039757 | 54777633 | 0.932 | HOXC4/ HOXC-AS1 | No | No | Yes |
| 12 | 15248516 | 16309290 | 0.755 | RERG/PTPRO | No | No | Yes |
| 3 | 47729665 | 49314960 | 0.628 | DALRD3/KLHDC8B/P4HTM/RBM6 | No | No | No |
| 1 | 76729016 | 79660928 | 0.618 | FUBP1/ADGRL4 | No | No | No |
| 10 | 19717815 | 22772115 | 0.551 | CASC10 | No | No | No |

**Figure 5.**
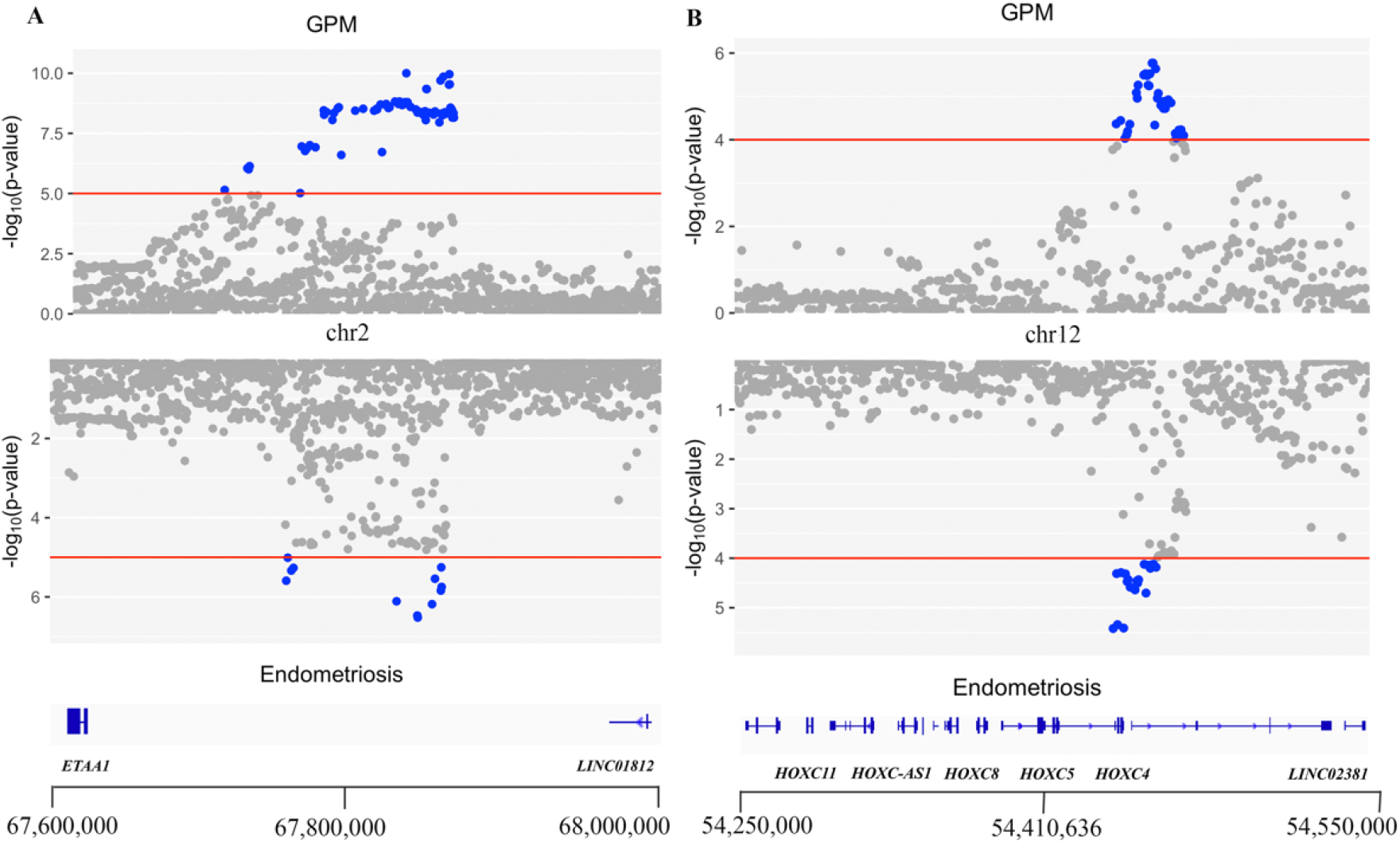
Mirror plot of first two GWAS-PW regions containing same causal variants for endometriosis and GPM. The left is the genomic locus near *ETAA1* on chromosome 2, while the right is the locus around *HOXC4* on chromosome 12.

### Gene mapping and functional annotation of shared risk loci

We performed a gene-based analysis using MAGMA to identify potential genes associated with both endometriosis and IBS or GPM. A total of 19 protein coding genes mapped to SNPs from the endometriosis and GPM cross-trait meta-analysis reached genome-wide significance (Supplementary Table 3). Only *FN1* passed the MAGMA analysis for SNPs from endometriosis and IBS meta-analysis.

To better understand the potential regulatory function of identified risk loci associated with both diseases, we mapped risk SNPs to cis-eQTLs in different tissues for genes up to 1MB on either side of the variant using FUMA. Lead SNPs in three loci (*RAB5B, ETAA1* and *HOXC*) from the cross-trait meta-analysis were found to impact gene expression in either blood or both digestive and reproductive tissues. In detail, we identified eQTLs for four genes in digestive tissues that contained SNPs associated with risk of endometriosis and IBS from the GWAS meta-analysis (Supplementary Table 4). Similarly, we identified eQTLs for 37 genes that contained SNPs associated with risk of endometriosis and GPM from the GWAS meta-analysis (Supplementary Table 4). Seven are expressed both in digestive and reproductive tissues. Using a more powerful blood eQTL dataset from the eQTLGen project we identified a total of 8 and 162 genes with eQTLs containing SNPs associated with risk of endometriosis and IBS, and GPM, respectively (Supplementary Table 4). Whilst 70% of genes with eQTLs in digestive and reproductive tissues were also in blood, indicating genetic regulation of gene expression can be shared across tissues, 12 were not identified in blood and may represent tissue specific effects. For example, the lead SNP, rs773111, in *RAB5B* locus shared by endometriosis and GPM is an eQTL for *RAB5B* expression in blood and for nearby *SUOX* gene expressed in blood, digestive, and reproductive tissues.

Combined annotation-dependent depletion (CADD) scores, which inform the deleterious effect of a SNP on protein function, were used to further understand the function of shared SNPs using FUMA. Our results showed that risk variants shared between endometriosis and IBS located in *WNT4* on chromosome 1 and *FN1* on chromosome 2 had at least one SNP with CADD score greater than 12.7 using positional gene mapping (Supplementary Table 5). Risk variants shared between endometriosis and GPM located in 33 different genes had at least one SNP with CADD score greater than 12.7 (Supplementary Table 5).

To investigate the shared functional mechanism of SNPs associated with both endometriosis and digestive disorders, we applied the Summary data-based Mendelian randomization (SMR) method. SMR integrates the GWAS summary statistics of disease with SNP-gene associations (eQTL), and the significant SMR association indicates that the SNP is causal for the disease as mediated through gene expression or has a pleiotropic effect on both disease and gene expression. We aimed to identify common associations between individual endometriosis and IBS and GPM SMR analyses. eQTL information in digestive and reproductive tissues in the GTEx project and in endometrium were used in this study. Our results identified 155 genes with normally significant SMR associations (*P*-SMR < 0.05; *P*-HEIDI > 0.05) for which variants were associated with risk of endometriosis and GPM and expression in digestive and reproductive tissues simultaneously (Supplementary Table 6). Of the 155 significant SMR associations, one variant (rs2344609), significantly associated with *CNGA4* expression, was significant in the cross-trait meta-analysis and another (rs9873183), associated with expression of *RNF123*, was in LD (r^2^ > 0.5) with significant SNPs in the cross-trait meta-analysis. When applied to endometriosis and IBS, we identified 91 genes with nominally significant SMR associations for variants associated with both diseases and expression in digestive and reproductive tissues (Supplementary Table 6). Of the 91 significant SMR associations, one variant (rs232877) associated with expression of *MYSM1* was in LD (r^2^ > 0.65) with the lead SNP of *TACSTD2* loci identified in the cross-trait meta-analysis, demonstrating that it may contribute to the risk of both endometriosis and IBS mediated through *MYSM1* expression.

Risk SNPs from the cross-trait metanalyses were also mapped to regulatory regions in reproductive and gastrointestinal tissues using EpiMap^49^. Of 477 SNPs significant in the endometriosis and GPM meta-analysis, 35 were located in predicted enhancers and nine of these enhancers were predicted in both reproductive and gastrointestinal tissues (Supplementary Table 7). Gene targets of these enhancers included 10 genes on chromosome 3 (*CYB561D2, GNAI2, HYAL3, NAT6, NPRL2, RBM5, RBM6, SEMA3F, SLC38A3, TUSC2*) and nine genes on chromosome 12 (*RERG, HOXC4, HOXC5, HOXC6, HOXC8, HOXC9, HOXC10, HOXC11, HOXC13*) including a cluster of HOXC genes (Supplementary Table 8). Five SNPs were located in predicter promoter regions for *RBM5* and *RBM6* on chromosome 3 and *HOXC-AS1, RERG* and *SUOX* on chromosome 12. Only one SNP significant in the endometriosis and IBS meta-analysis was located in a regulatory region, a predicted enhancer and promoter region in *FN1* on chromosome 2 in colon and esophagus (Supplementary Table 7).

In total, 218 genes were mapped to 24 shared risk loci using at least one annotation method above (Supplementary Table 9) and 37 genes had evidence from at least two methods (Table 5).

**Table 5.**
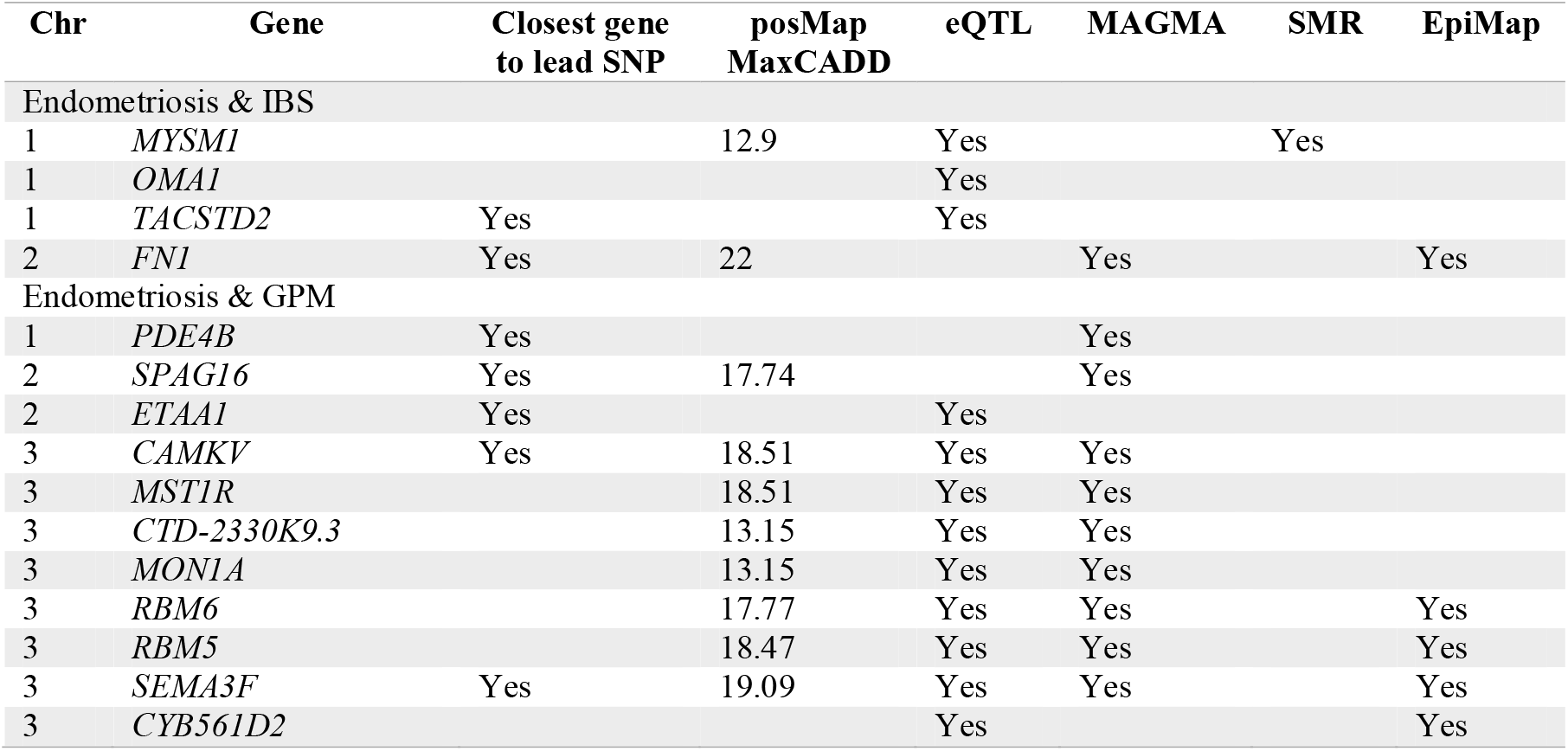
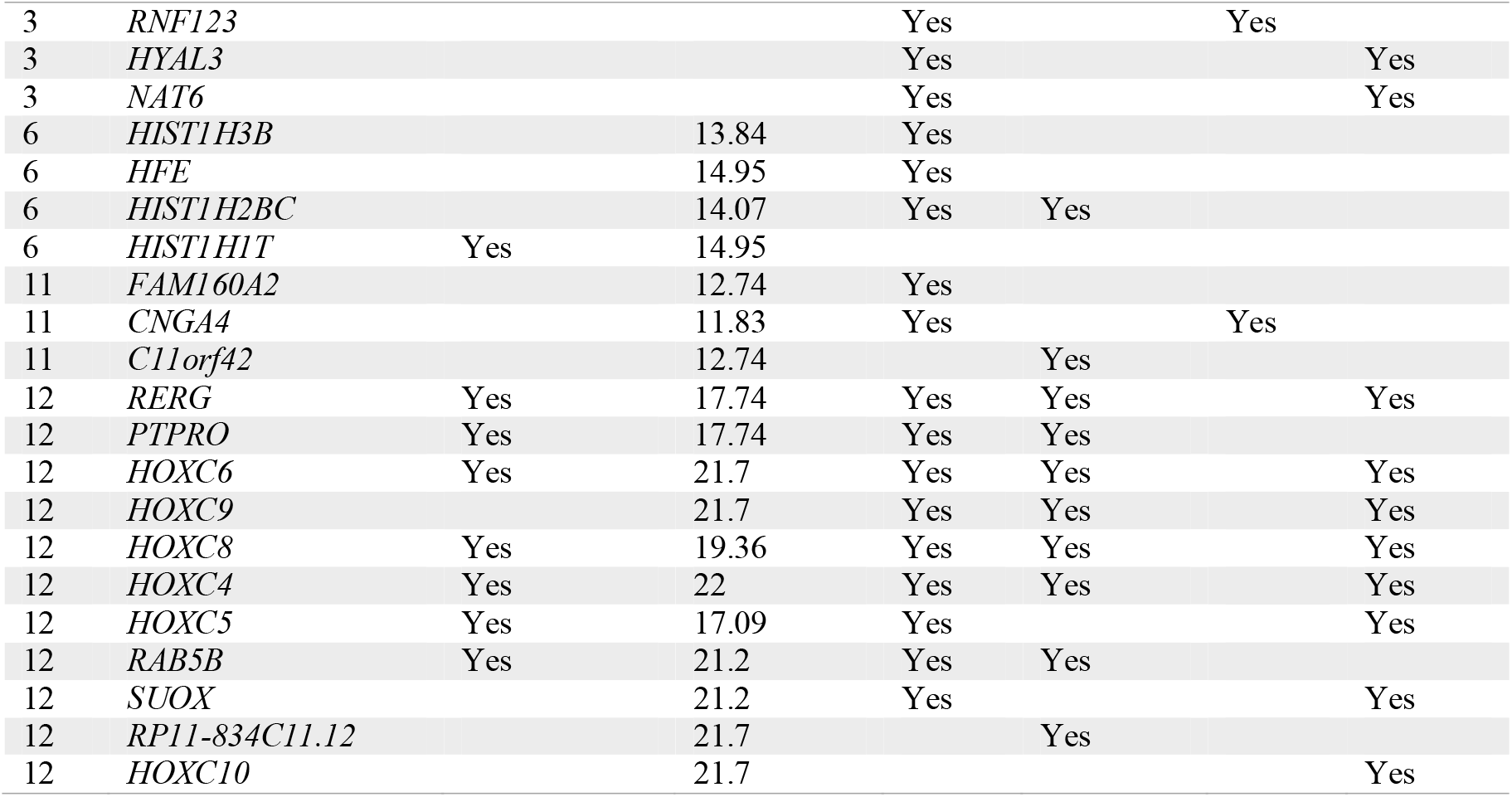
Functionally mapped genes for significant risk loci identified in the cross-trait meta-analysis of endometriosis and gastrointestinal disorders.

### Pathways enriched for genes in shared risk loci

The MAGMA gene-set analysis, which uses the full distribution of input SNP p-values to identify whether curated gene sets and GO terms from MsigDB were significantly associated with both diseases, found no gene set remained significant after multiple testing. We instead used another pathway enrichment approach, GENE2FUNC, to test whether 204 prioritized genes (eQTL, CADD, MAGMA, SMR) linked to endometriosis and GPM were enriched in pre-defined pathways based on their gene expression. As a result, a total number of five KEGG gene sets passed adjusted p-value threshold of *P* < 0.05, including “leukocyte transendothelial migration”, “oxidative phosphorylation”, “epithelial cell signalling in helicobacter pylori infection”, and “chemokine_signaling pathway” (Supplementary Table 10). Genes were also enriched in 40 GWAS Catalog reported gene sets, including fat distribution, BMI, intelligence, and depression (Supplementary Table 10). The 14 genes linked to endometriosis and IBS were enriched in two KEGG pathways including “pathway in cancer” and “ECM receptor interaction” and 17 GWAS Catalog gene sets (Supplementary Table 11).

### Additional phenotypes associated with shared risk loci

Given the nature of SNP pleiotropy, we further investigated whether there were previously reported trait associations with SNPs related to risk of both endometriosis and gastrointestinal diseases through PhenoScanner and GWAS Catalog. Interestingly, as shown from the results in Supplementary Table 12, our results identified that fat and estrogen related traits (BMI, body fat percentage, waist circumference, hip circumference, WHR, weight, age at first birth and age at menarche) are associated with six (*WNT4, SEMA3F, HIST1H2BC, RERG, RAB5B, HOXC4/5/6*) of twelve regions shared between endometriosis and GPM, which consistently supported that those identified regions might contribute to the risk of both endometriosis and GPM through the dysregulation of estrogen and inflammation. The precise interaction mechanism of estrogen and fat related traits with endometriosis and GPM is unknown and requires further investigation.

### Potential for drug repositioning

The online Open-target drug platform was used to assess if any of the genes linked to both endometriosis and the GI disorders were potential drug targets (https://www.targetvalidation.org). A total of 218 unique genes with evidence from gene mapping and functional annotation (Supplementary Table 9) were used to search for known endometriosis and gastrointestinal disorder (GORD, PUD, IBS) drug targets. One gene, *CCKBR* encoded a protein that was targeted by two drugs Proglumide (ATC code: A02BX06) and Netzepide (NCT01298999 and NCT02597712) for the clinical treatment of GORD and PUD. In addition, *PDE4B* with its encoded protein cAMP-specific 3’,5’-cyclic phosphodiesterase 4B being targeted by Pentoxifylline, acts as an inhibitor targeting the immune system and has been clinically trialled for the treatment of both endometriosis (phase III) and IBS (Phase IV) separately (Table 6). Compared with IBS which has multiple sources of evidence to support the promising treatment effect of Pentoxifylline^51 52^, there is limited evidence on whether Pentoxifylline impacts endometriosis related pain reduction ^53^. Notably, when not restricted to the aforementioned gene set, 34 genes with encoded proteins were targets of both endometriosis and IBS/GORD/PUD drugs (Supplementary Table 13). This study may provide novel insights and evidence for the further investigation of therapeutic targets for both endometriosis and GI traits.

**Table 6.**
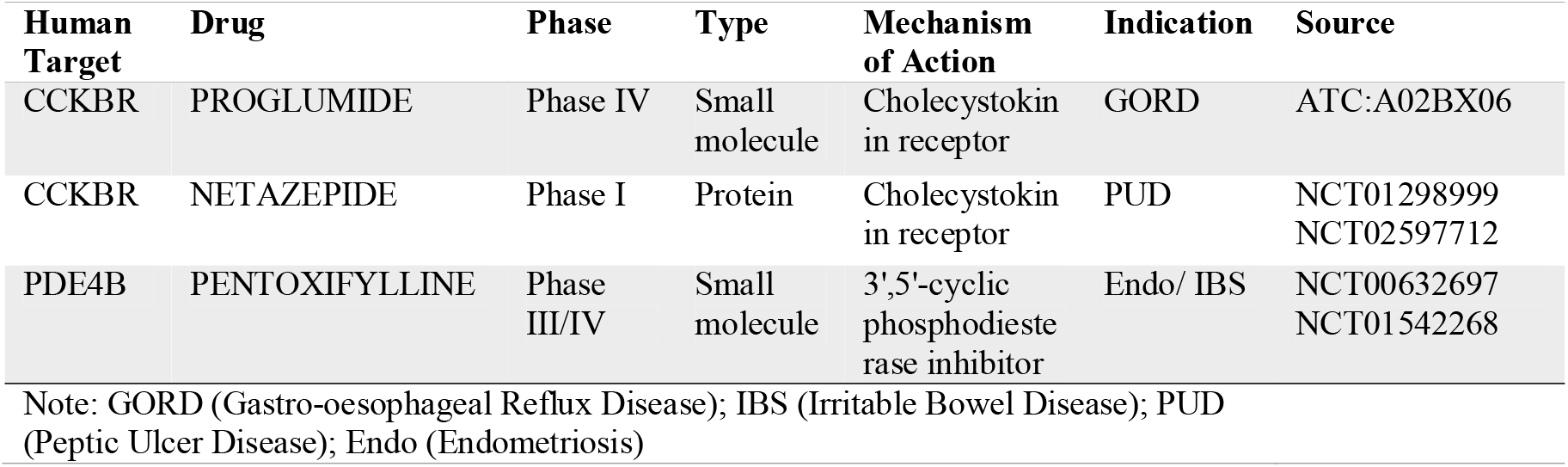
Existing drug targets for endometriosis and gastrointestinal disorders.

| Human Target | Drug | Phase | Type | Mechanism of Action | Indication | Source |
| --- | --- | --- | --- | --- | --- | --- |
| CCKBR | PROGLUMIDE | Phase IV | Small molecule | Cholecystokin in receptor | GORD | ATC:A02BX06 |
| CCKBR | NETAZEPIDE | Phase I | Protein | Cholecystokin in receptor | PUD | NCT01298999<br>NCT02597712 |
| PDE4B | PENTOXIFYLLINE | Phase III/IV | Small molecule | 3',5'-cyclic phosphodiesterase inhibitor | Endo/ IBS | NCT00632697<br>NCT01542268 |

Note: GORD (Gastro-oesophageal Reflux Disease); IBS (Irritable Bowel Disease); PUD (Peptic Ulcer Disease); Endo (Endometriosis)

### Insights from medication use in both diseases

Identified comorbid relationships between endometriosis and gastrointestinal disorders and shared symptomology as well as shared genetic components also raise questions around the potential effects of medication use on disease aetiology and management. We investigated prescription medication use in women by endometriosis status using data from the PBS records for both the 1973-78 and the 1989-95 ALSWH cohorts. Interestingly, drugs for peptic ulcer and gastro-oesophageal reflux were within the top 10 most frequently used drugs in both cohorts and such drug usage rate was significantly higher in women with endometriosis than those without in 1989-95 ALSWH cohort after multiple testing (Supplementary Table 14), further evidence of the likely co-occurrence of the diseases and disease symptoms. Consistently, using age-matched medication data of unrelated European women within the UK biobank revealed that in addition to the expected hormonal therapies and NSAIDs, up to seven medications for treatments of GORD, PUD and IBS were also significantly higher in women with endometriosis compared with women without (Table 7 & Supplementary Table 15). More interestingly, when comparing medication usage between women with and without gastrointestinal disorders in the UK biobank, we also found a significantly higher use of hormone therapies among IBS, GORD, and PUD, but not for IBD (Table 7 & Supplementary Table 16), which is consistent with genetic results in this study. We next searched for the target genes of those drugs used for GI disorders identified above, and found that *SLC22A1*, one of their transporters, has also been identified by our cross-trait meta-analysis and subsequent functional mapping (Supplementary Table 4 & Supplementary Table 9).

**Table 7.** Comparison of medication usage in UKB unrelated European women with and without a diagnosis of endometriosis or gastrointestinal disorders.

| Medication | Diagnoses |  | Controls |  | P.Value | BF.P.value | Description |
| --- | --- | --- | --- | --- | --- | --- | --- |
|  | Med | No | Med | No |  |  |  |
| Endometriosis |  |  |  |  |  |  |  |
| Omeprazole | 540 | 4852 | 5521 | 86823 | 1.49E-28 | 1.73E-25 | GORD/PUD Treatment |
| Laxatives | 353 | 5039 | 3749 | 88595 | 1.52E-16 | 1.76E-13 | Constipation Treatment |
| Lansoprazole | 266 | 5126 | 2881 | 89463 | 6.96E-12 | 8.05E-09 | GORD/PUD treatment |
| Mebeverine | 73 | 5319 | 558 | 91786 | 3.84E-09 | 4.45E-06 | IBS Treatment |
| Ranitidine | 163 | 5229 | 1765 | 90579 | 1.06E-07 | 0.0001 | GORD/PUD treatment |
| Senna | 30 | 5362 | 176 | 92168 | 1.25E-06 | 0.0015 | Constipation Treatment |
| Esomeprazole | 33 | 5359 | 242 | 92102 | 2.88E-05 | 0.0334 | GORD/PUD treatment |
| Paracetamol | 1914 | 3478 | 25462 | 66882 | 8.16E-35 | 9.45E-32 | Painkiller |
| Premarin | 132 | 5260 | 595 | 91749 | 1.49E-33 | 1.73E-30 | Hormone Therapy |
| Irritable Bowel Disease |  |  |  |  |  |  |  |
| Vagifem | 200 | 15679 | 758 | 126274 | 2.29E-18 | 2.91E-15 | Hormone Therapy |
| Premarin | 185 | 15694 | 891 | 126141 | 2.31E-09 | 2.94E-06 | Hormone Therapy |
| Estraderm | 80 | 15799 | 291 | 126741 | 6.54E-09 | 8.31E-06 | Hormone Therapy |
| Ovestin | 51 | 15828 | 151 | 126881 | 1.59E-08 | 2.03E-05 | Hormone Therapy |
| Estradiol product | 99 | 15780 | 430 | 126602 | 2.33E-07 | 0.0003 | Hormone Therapy |
| Evorel 25 patch | 82 | 15797 | 346 | 126686 | 9.59E-07 | 0.0012 | Hormone Therapy |
| Conjugated oestrogens | 18 | 15861 | 29 | 127003 | 1.22E-06 | 0.0016 | Hormone Therapy |
| Climaval 1mg tablet | 65 | 15814 | 258 | 126774 | 2.08E-06 | 0.0026 | Hormone Therapy |
| Estriol product | 37 | 15842 | 114 | 126918 | 3.17E-06 | 0.0040 | Hormone Therapy |
| Omeprazole | 2242 | 13637 | 4209 | 122823 | 0 | 0 | GI disorder treatment |
| **Gastro-oesophageal Reflux Disease** |  |  |  |  |  |  |  |
| Estradiol product | 134 | 22248 | 317 | 89211 | 8.50E-07 | 0.0010 | Hormone Therapy |
| Estraderm | 94 | 22288 | 217 | 89311 | 1.81E-05 | 0.0207 | Hormone Therapy |
| Premarin | 233 | 22149 | 672 | 88856 | 2.44E-05 | 0.0279 | Hormone Therapy |
| Lansoprazole | 3705 | 18677 | 1640 | 87888 | 0 | 0 | GI disorder treatment |
| Omeprazole | 6948 | 15434 | 3335 | 86193 | 0 | 0 | GI disorder treatment |
| Ranitidine | 1596 | 20786 | 1081 | 88447 | 0 | 0 | GI disorder treatment |
| Gaviscon liquid | 714 | 21668 | 237 | 89291 | 1.01E-295 | 1.15E-292 | GI disorder treatment |
| Paracetamol | 7900 | 14482 | 21440 | 68088 | 1.92E-249 | 2.20E-246 | Painkiller |
| Esomeprazole | 434 | 21948 | 99 | 89429 | 3.35E-205 | 3.83E-202 | GI disorder treatment |
| **Peptic Ulcer Disease** |  |  |  |  |  |  |  |
| Estraderm | 35 | 5173 | 205 | 72707 | 1.26E-05 | 0.0135 | Hormone Therapy |
| Omeprazole | 1375 | 3833 | 2821 | 70091 | 0 | 0 | GI disorder treatment |
| Lansoprazole | 711 | 4497 | 1426 | 71486 | 2.34E-306 | 2.49E-303 | GI disorder treatment |
| Ranitidine | 351 | 4857 | 942 | 71970 | 3.67E-118 | 3.91E-115 | GI disorder treatment |
| Paracetamol | 1963 | 3245 | 17305 | 55607 | 1.74E-103 | 1.86E-100 | Painkiller |
| Esomeprazole | 107 | 5101 | 88 | 72824 | 1.62E-72 | 1.73E-69 | GI disorder treatment |
| Tramadol | 217 | 4991 | 665 | 72247 | 1.10E-64 | 1.17E-61 | Opioid |
| Co-codamol | 348 | 4860 | 1608 | 71304 | 4.27E-64 | 4.55E-61 | Opioid |
| Amitriptyline | 326 | 4882 | 1477 | 71435 | 1.03E-61 | 1.10E-58 | Antidepressants |

## Discussion

### Summary findings of this study

This is the first study to comprehensively illustrate the link between endometriosis and gastrointestinal disorders which both affect a large proportion of people worldwide, using large-scale multi-dimensional data including clinical, genetic, and pharmaceutical datasets. We confirmed both a bidirectional epidemiological association and a shared genetic basis of endometriosis with each of the three GI traits (IBS, GORD, and PUD). Evidence of medication use further supported the co-occurrence and shared disease aetiology of these conditions but also highlights the potential for drug repositioning and caution around drug contraindications in clinical practice.

### Evidence for comorbid relationships

Endometriosis and IBS, two of the leading causes of chronic pelvic pain, are often misdiagnosed in clinics due to non-specific symptoms^54^. Several epidemiological studies indicated women with endometriosis have an increased risk of a diagnosis of IBS^55 56^. For example, one nationwide UK case-control study demonstrated women diagnosed with endometriosis were two and half times more likely to receive a new diagnosis of IBS when compared with controls (OR = 2.5, 95% CI 2.2 to 2.8)^54^. Few studies have investigated if women with IBS are more likely to be diagnosed with endometriosis. Our analysis using diagnoses reported in UKB supported the epidemiological association between endometriosis and IBS. We identified a bidirectional association between endometriosis and all four gastrointestinal disorders. Of those associations, endometriosis showed a stronger relationship with IBS and GORD when compared with PUD and IBD. Therefore, in addition to the clinically reported shared symptomology, this study provides further evidence of a complex phenotypic association between endometriosis and gastrointestinal disorders.

### Evidence of shared genetic aetiology and potential drug candidates

A previous study investigating the shared genetic basis between endometriosis and depression has implicated gastric mucosa abnormalities in this casual pathway^29^. Adewuyi et al. reported a strong genetic correlation between endometriosis and GORD using two early published GWAS summary datasets in the UKB^29^ however, other common GI disorders such as IBS were not measured. Consistent with previous reports, our study, which included one third more individuals from the UKB, also identified a significant genetic correlation between endometriosis and GORD. In addition, our study identified novel genetic correlations between endometriosis and IBS and PUD, but not for IBD. This is in line with known genetic differences between IBD and the other three GI disorders identified using both partitioned SNP-based heritability analysis and bivariate LDSC analysis^30^. When using a more powerful phenotype GPM, a combination of diagnosis of PUD and/or GORD as two acid-related diseases that share treatment therapy in clinical practice, we found a stronger genetic correlation with endometriosis. Many observational studies are subject to the confounding effects of environment and lifestyle factors. In contrast, our analysis based on genotype-level data are unlikely to suffer from such methodological bias. Therefore, the estimated one-fifth of the genetic contribution to endometriosis, which is shared with the genetic contribution to IBS or GPM, may partly explain the significant comorbid relationship between the diseases.

In addition to a genetic correlation between endometriosis and IBS and GPM we identified a bidirectional causal relationship between genetic risk of IBS and endometriosis and a unidirectional causal relationship between GPM and endometriosis. The MR framework is a powerful and cost-effective method for inferring causality due to its advantage that genetic alleles are determined at conception, so that MR results are free from potential environmental confounders (eg. medication usage) and bias which are often found in observational studies^57^. In this study, the identification of bidirectional causality (vertical pleiotropy) between endometriosis and IBS suggests that the increased risks of endometriosis in IBS patients and vice versa are, in part, mediated by the genetic liability to the other disease. The estimated causal relationship between traits can also be affected by violations of MR assumptions which occur when genetic variants are also associated with confounding factors or influence the outcome through a confounding factor. A strength of our study is that we conducted the mtCOJO conditional analysis which takes into account the correlation between traits given the previously reported complexity among GI disorders^30^, and the significant bidirectional relationship between IBS and GPM in this study using both standard and reverse MR analysis (Figure 3). The MR results remained similar after conditioning on GPM, indicating that the bidirectional causal relationship between endometriosis and IBS at the genetic level were not driven by their relationship with GPM.

Moreover, the subsequent identification of shared genomic loci using cross-trait meta-analysis and colocalization approaches further provides clues as to the possible biological mechanisms and specific pathways driving the causal relationships between the different gastrointestinal disorders and endometriosis. For example, both *TACSTD2* on chromosome 1 and *FN1* on chromosome 2 shared by endometriosis and IBS are involved in various cellular processes including cell proliferation, motility, invasion and migration ^58,59^. *TACSTD2* is a novel region that has not been implicated in IBS or endometriosis GWAS analyses previously. The intracellular calcium signal transducer *TACSTD2* was reported to be overexpressed in endometrioid-type endometrial carcinoma and gastrointestinal cancers ^60 61^. Another shared locus *FN1* has been previously linked to the risk of endometriosis at a genome-wide significance threshold of *P* < 5e-8 in both the latest GWAS study^62^ and a recent Greek population-based GWAS study^63^. Despite *FN1* not having been identified in IBS GWAS meta-analyses, there are studies demonstrating a significant down-regulation of expression of *FN1* in IBS patients compared with controls, which may be responsible for the increased mucosal permeability and visceral hypersensitivity of IBS through its mediation in barrier dysfunction ^64 65^. Functional annotations in this study also suggest a possible pathogenic effect of these two regions, providing evidence that the lead SNP (rs6661808) at the *TASCTD2* locus is an eQTL in both blood and digestive tissues, while the lead SNP in the *FN1* region was within a predicted enhancer or promoter in digestive tissues and had a CADD score over 12.7.

Three genomic loci shared by endometriosis and GPM were identified in this study using both cross-trait meta-analysis and GWAS-PW, including *ETAA1* on chromosome 2, *HOXC* and *RERG* on chromosome 12. *ETAA1* is an activator of ATR kinase which plays a key role in protecting the genome against both intrinsic replication problems and substantial extrinsic DNA damage^66^, while the other two regions are closely associated with receptor signalling and estrogen metabolism ^67-70^. The *ETAA1* locus has been previously reported to be associated with risk of both traits at genome-wide significance level^30 62^. Functional annotation of independent SNPs in the *HOXC* region suggest that these variants impact regulatory elements in both digestive and reproductive tissues and regulate expression of several HOXC genes (*HOXC4, HOXC5, HOXC6, HOXC8, HOXC9, HOXC10, HOXC-AS1)*.

Altered expression of the HOXC cluster has been found in ectopic and eutopic tissues from endometriosis patients vs. control^71-73^, as well as in gastrointestinal disorders such as ulcerative colitis, colorectal cancer and gastric cancer^74-77^. Even though *RERG*, and nearby *PTPRO*, were not implicated in any GWAS studies of the separate traits, P-values for the lead SNP were close to genome-wide significance (*P*_Endometriosis_ = 9.24E-06, *P*_GPM_ = 8.80E-07) suggesting the increased power from combining the traits was able to identify a novel risk locus for both diseases. Moreover, experiments have shown that the upregulated ERβ expression and attenuated ERα expression in endometriosis lesions indicated that insufficient expression of *PTPRO* may be involved in the progression of endometriosis^78^. Research findings on the association between estrogen and GORD and PUD have been contradictory^79-84^, evidenced by some studies reporting a higher prevalence rate of GORD and PUD in men compared with women before the age of menopause^81 85^ and others reporting a positive correlation between GORD symptoms and postmenopausal hormone therapy^86^. In this study, we provided additional evidence that this estrogen related loci on chromosome 12 may be involved in the progression of both endometriosis and GORD and PUD however, the underlying molecular mechanism remains unclear.

In addition to the five highlighted regions shared by endometriosis and IBS/GPM, another six regions (*SEMA3F, SPAG16, HIST1H2BC, RAB5B, CCKBR* and *PDE4B*) identified by cross-trait meta-analysis and at least two functional annotation analyses also implicate potential pathways that are associated with the two traits. With the exception of *HIST1H2BC* and *SPAG16* loci, the remaining four loci have been implicated in previous GORD or PUD GWAS studies ^30 87^. The lead SNPs in these six regions are all nominally significantly associated with endometriosis and have not been linked to endometriosis in previous GWAS studies. They may represent novel target genes or pathways involved in endometriosis progression. For example, *HIST1H2BC, SPAG16, SEMA3F* and *RAB5B* themselves or genes nearby those regions may be associated with endometriosis by either being the estradiol responsive gene, or related to steroid hormone treatment and metabolism, and further regulate the proliferation of endometrial stromal cells^88-95^.

### Evidence for the potential of drug repositioning in clinics

To verify whether the 218 candidate genes identified have any implications in the clinic, we searched the online drug target databases and identified *CCKBR* and *PDE4B* with their encoded proteins as drug targets. While the former is currently used for treatment of PUD and GORD, the latter has been clinically trialled for both IBS and endometriosis. *PDE4B* is mainly present in immune and epithelial cells and has a role modulating inflammation and epithelial integrity ^96^ while *CCKBR* encodes a G-protein coupled receptor for gastrin and cholecystokinin. Even though *CCKBR* has not been targeted for treatment of endometriosis, a recent study has demonstrated that reduction of gastrin is associated with inactivation of CCKBR/ERK/P65 signalling in estrogen receptor positive breast cancer cells, and lower expression of gastrin and *CCKBR* was correlated to worse prognosis in breast cancer^97^. Therefore, the involvement of gastrin and *CCKBR* in estrogen metabolism may implicate this gene as a potential drug target for endometriosis. Moreover, considering the low quality of current evidence for Pentoxifylline which was used to treat endometriosis by targeting *PDE4B* encoded protein^53^, this study provides more evidence and novel insight for the further investigation of *PDE4B* for the purpose of treating both endometriosis and GI disorders.

### Evidence for associations with medication usage

In the present study, our ability to link diagnoses with drug usage further revealed overlap in medication use between endometriosis and GI disorders, providing novel insights for disease aetiology and management in clinics. The identification of a higher use of drugs for IBS, GORD and PUD in women diagnosed with endometriosis as well as the higher use of hormone therapies in women diagnosed with IBS, GORD and PUD but not for IBD, strongly supports the coexistence and potential manifestations of underlying pathophysiological and genetic correlations between the diseases. Whilst not unexpected, the frequent use of NSAIDs by women with endometriosis highlights potential confounding from contraindicated therapies as the frequent use of NSAIDs during endometriosis treatment is a well-known risk factor for PUD by destroying the mucus layer in the digestive tract^15^. However, the genetic correlation between the diseases suggests the relationship is not driven by a consequence of medical therapies alone. Additionally, we also identified that the frequency of other treatments for gastrointestinal disorders such as omeprazole and laxatives was also significantly higher in women with endometriosis than women without. One of the gene targets of omeprazole, *ATP4A*, is a member of the ATPase family responsible for oxidative phosphorylation. Evidence from the cross-trait meta-analysis and functional annotation methods highlighted *ATP6V0E1* and *ATP6V0E2* as potential target genes involved in both endometriosis and GPM. *ATP6V0E1* and *ATP6V0E2* also belong to ATPase family suggesting targeting this pathway may have effects on both GI disorders and endometriosis. Other evidence towards the potential of drug repositioning includes that visceral sensitivity and chronic low-grade inflammatory state have been key characteristics in both IBS and endometriosis^98^ and therapies targeted at relieving pain in IBS can also relieve pain during menstruation^99^. Similarly, a New Zealand based study reported that women with IBS and concurrent endometriosis had a significantly higher response rate to a low FODMAP diet, one therapy for IBS, than those IBS patients with no known endometriosis^100^.

### Clinical implications

At least three types of clinical implication can be drawn from this study. First, regarding diagnosis for both endometriosis and GI disorders, shared aetiology suggests joint or alternative diagnoses should considered for patients presenting symptoms related to either disease. Second, evidence from medication use suggest caution around contraindications for some drugs, as NASIDs, often used for endometriosis, are a well-known risk factor for PUD. Therefore, it may be worthwhile for clinicians to consider potential contraindications when prescribing NASIDs for female patients presenting with symptoms shared between the diseases such as abnormal pain, bloating or constipation, etc. Third, support for *PDE4B* as a shared drug target for the treatment of both IBS and endometriosis, suggest comorbidity of endometriosis and IBS should be considered in the design and recruitment for clinical trials.

### Strengths and limitations

One major strength of the current study is the use of large-scale population data, genetic and medication usage data for both gastrointestinal disorders and endometriosis, to comprehensively illustrate the association between these two disorders. Integrating these datasets provides more convincing evidence for the association of diseases, as well as important clinical implications. Despite these interesting findings, we acknowledge serval limitations. First, endometriosis is a highly heterogeneous condition with variation in lesion location and grade^101^. Similarly, the four GI phenotypes derived from UKB may also introduce further heterogeneity^30^. It is not clear if certain subtypes of endometriosis share more genetic risk factors with digestive disorders. Therefore, the associations identified in this study should be validated in larger endometriosis datasets with more detailed phenotype information when these are available. Second, as mentioned in Wu et al.^30^, the existence of co-reporting of some diagnoses, including two gastrointestinal disorders, may bias the association with specific digestive disorders. However, the sensitivity tests carried out in the study, which excluded those individuals with more than one diagnosis, demonstrated that the co-existence did not impact conclusions. We concluded that GWAS summary statistics for IBS and GPM phenotypes are robust, and identified different genetic risk factors shared between endometriosis and IBS and GPM. Third, compared with the other four phenotypes, there are fewer IBD cases available in UKB, which may limit the power for both genetic and epidemiological analyses. Results for the IBD GWAS were highly consistent with previously published GWAS^30^ suggesting results in this study are robust.

## Conclusions

This study comprehensively assesses the observational, genetic, and pharmaceutical usage associations between endometriosis and gastrointestinal disorders using various statistical approaches and multidimensional large-scale datasets. We provide strong evidence for the shared aetiology of endometriosis and digestive disorders and highlight target genes and pathways contributing to the shared aetiology. The results suggest potential targets for treatment, considerations for disease management and caution around contraindications for some drugs. The clinical implications could facilitate better clinical outcomes for women with both endometriosis and gastrointestinal diseases.

## Data Availability

This study includes no data deposited in external repositories.

## Data Availability

This study includes no data deposited in external repositories.

## Author Contributions

F.Y, S.M and G.W.M designed the study with input from the other authors. G.W.M, S.M, Y.W, R.H and G.D.M coordinated data collection, quality control of data, data management and analysis of the original datasets. F.Y, S.M, R.H and Y.W ran additional quality control and filtering of datasets. Data analysis was performed by F.Y which was interpreted by all authors. F.Y, S.M, and G.W.M drafted the report with input from all other authors. The final manuscript has been critically revised and approved by all authors.

## Acknowledgements

This research has been conducted using the UK Biobank Resource under Application Number 54861 and 12505. Summary statistics from the endometriosis GWAS used in this study contain data from 23andMe. We would like to thank the research participants and employees of 23andMe, Inc. for making this work possible.

The research on which this paper is partly based was conducted as part of the Australian Longitudinal Study on Women’s Health by the University of Queensland and The University of Newcastle. We are grateful to the Australian Government Department of Health and Aged Care for funding and to the women who provided the survey data.

We acknowledge the Department of Health and Medicare Australia for providing MBS and PBS data and the Australian Institute of Health and Welfare (AIHW) as the integrating authority. We also acknowledge the following:

- Centre for Health Record Linkage (CHeReL), NSW Ministry of Health and ACT Health, for the NSW Admitted Patients Data Collection, and the ACT Admitted Patient Care Data Collections.
- Queensland Health, including the Statistical Services Branch, for the Qld Hospital Admitted Patient Data Collection.
- Department of Health Western Australia, including the Data Linkage Branch, and the WA Hospital Morbidity Data Collection.
- SA NT Datalink, and SA Department for Health and Wellbeing and Northern Territory Department of Health, for the SA Public Hospital Separations and NT Public Hospital Inpatient Activity Data Collections.
- Tasmanian Data Linkage Unit, and the Department of Health, Tasmania, for the Public Hospital Admitted Patient Episodes Data Collection.
- Victorian Department of Health as the source of the Victorian Admitted Episodes Dataset, and the Centre for Victorian Data Linkage (Victorian Department of Health) for the provision of data linkage.

## The International Endometriosis Genetics Consortium (IEGC)

The following are members of the International Endogene Consortium (IEC): Yadav Sapkota, Valgerdur Steinthorsdottir, Andrew P. Morris, Amelie Fassbender, Nilufer Rahmioglu, Immaculata De Vivo, Julie E. Buring, Futao Zhang, Todd L. Edwards, Sarah Jones, Dorien O, Daniëlle Peterse, Kathryn M. Rexrode, Paul M. Ridker, Andrew J. Schork, Stuart MacGregor, Nicholas G. Martin, Christian M. Becker, Sosuke Adachi, Kosuke Yoshihara, Takayuki Enomoto, Atsushi Takahashi, Yoichiro Kamatani, Koichi Matsuda, Michiaki Kubo, Gudmar Thorleifsson, Reynir T. Geirsson, Unnur Thorsteinsdottir, Leanne M. Wallace, iPSYCH-SSI-Broad Groupw, Jian Yang, Digna R. Velez Edwards, Mette Nyegaard, Siew-Kee Low, Krina T. Zondervan, Stacey A. Missmer, Thomas D’Hooghe, Grant W. Montgomery, Daniel I. Chasman, Kari Stefansson, Joyce Y. Tung, and Dale R. Nyholt

