## Supplementary Figure 1 for "Evidence of shared genetic factors in the aetiology of gastrointestinal disorders and endometriosis and clinical implications for disease management"

**Supplementary Figure 1.** Genomic loci shared by endometriosis and IBS/GPM using GWAS-PW analyses (PPA3 > 0.5). A-C represents shared loci between endometriosis and IBS while D-G represents shard loci between endometriosis and GPM. A, locus around *FN1* on chromosome 2; B, locus near to *TACSTD2* on chromosome 1; C, locus around *HLA-C* on chromosome 6; D, locus near to *RERG* on chromosome 12; E, locus around *RBM6* on chromosome 3; F, locus near to *ADGRL4* on chromosome 1; G, locus around *CASC10* on chromosome 10.

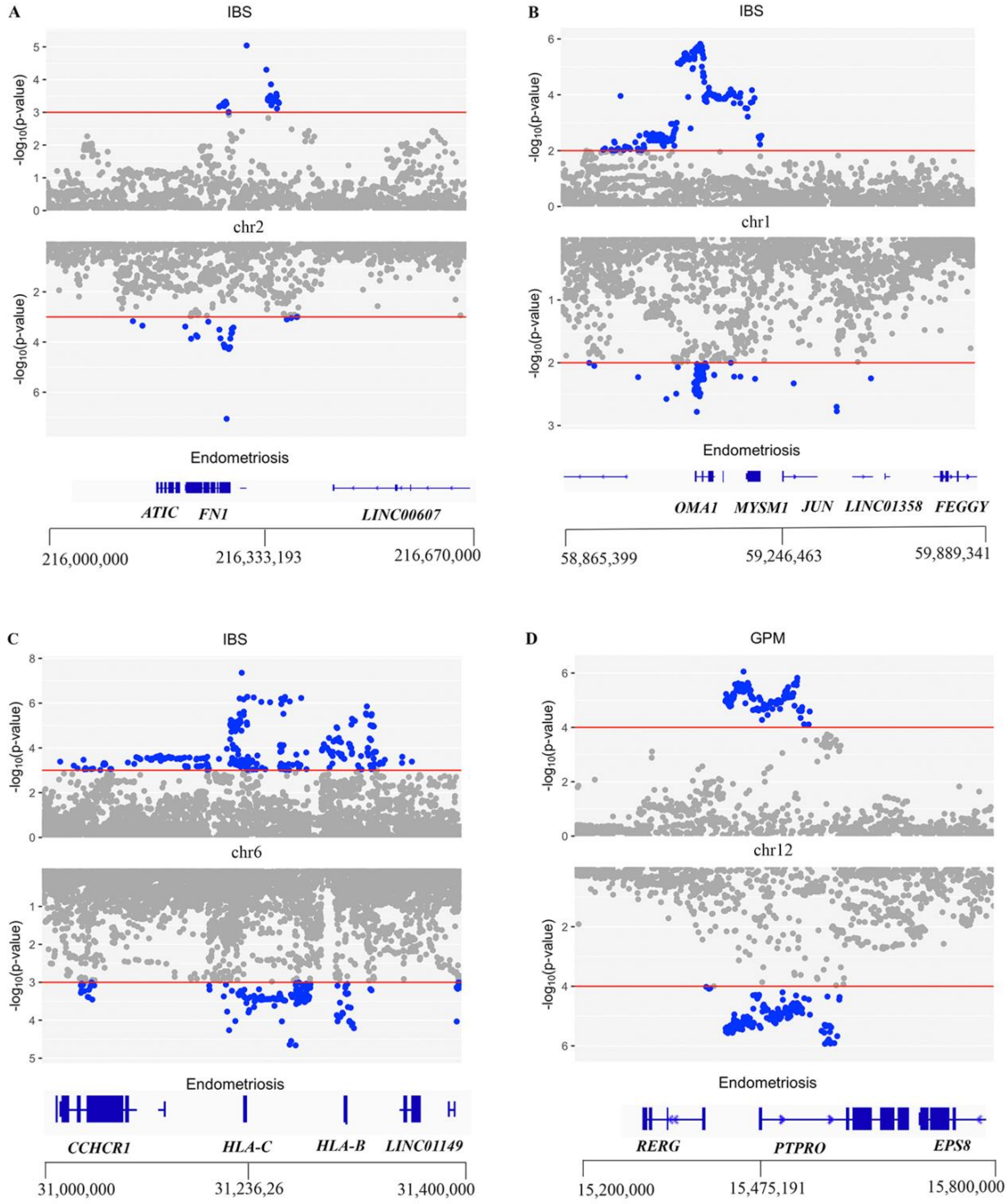

E

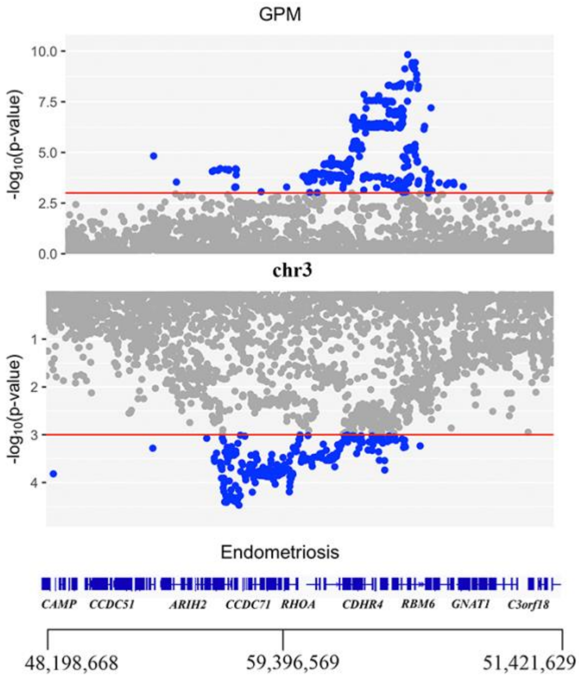
GPM
-log10(p-value)
chr3
Endometriosis
CAMP
CCDC51
ARIH2
CCDC71
RHOA
CDHR4
RBM6
GNAT1
C3orf18
48,198,668
59,396,569
51,421,629

F

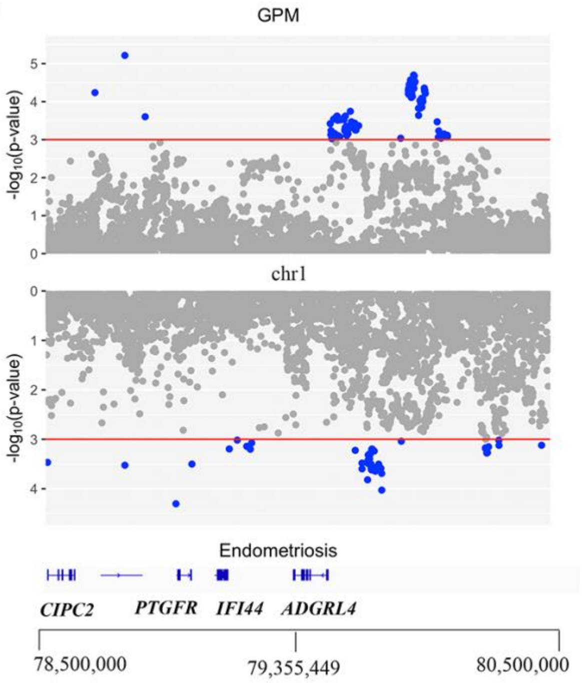
GPM
-log10(p-value)
chr1
Endometriosis
CIPC2
PTGFR
IFI44
ADGRL4
78,500,000
79,355,449
80,500,000

G

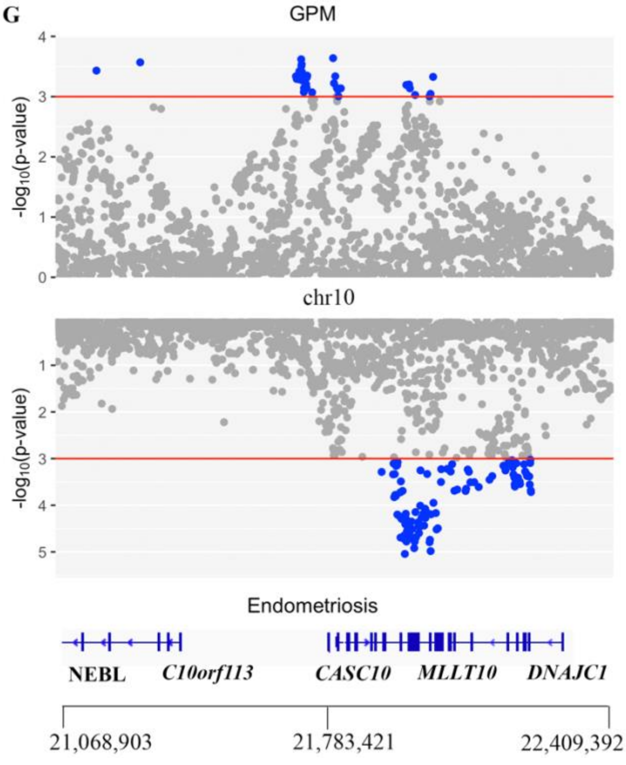
GPM
-log10(p-value)
chr10
Endometriosis
NEBL
C10orf113
CASC10
MLLT10
DNAJC1
21,068,903
21,783,421
22,409,392
